# Rewiring of cortical glucose metabolism fuels human brain cancer growth

**DOI:** 10.1101/2023.10.24.23297489

**Authors:** Andrew J. Scott, Anjali Mittal, Baharan Meghdadi, Sravya Palavalasa, Abhinav Achreja, Alexandra O’Brien, Ayesha U. Kothari, Weihua Zhou, Jie Xu, Angelica Lin, Kari Wilder-Romans, Donna M. Edwards, Zhe Wu, Jiane Feng, Anthony C. Andren, Li Zhang, Vijay Tarnal, Kimberly A. Redic, Nathan Qi, Joshua Fischer, Ethan Yang, Michael S. Regan, Sylwia A Stopka, Gerard Baquer, Theodore S. Lawrence, Sriram Venneti, Nathalie Y. R. Agar, Costas A. Lyssiotis, Wajd N. Al-Holou, Deepak Nagrath, Daniel R. Wahl

**Affiliations:** Department of Radiation Oncology, University of Michigan, Ann Arbor, MI, USA; Rogel Cancer Center, University of Michigan, Ann Arbor, MI, USA; Laboratory for Systems Biology of Human Diseases, University of Michigan, Ann Arbor, MI, USA; Biointerfaces Institute, University of Michigan, Ann Arbor, MI, USA; Department of Chemical Engineering, University of Michigan, Ann Arbor, MI, USA; Department of Biomedical Engineering, University of Michigan, Ann Arbor, MI, USA; Department of Molecular and Integrative Physiology, University of Michigan, Ann Arbor, MI, USA; Department of Anesthesiology, University of Michigan, Ann Arbor, MI, USA; Department of Pharmacy Services, Michigan Medicine, University of Michigan, Ann Arbor, MI, USA; Bruker, Billerica, MA, USA; Department of Neurosurgery, Brigham and Women’s Hospital, Harvard Medical School, Boston, MA, USA; Department of Pathology, University of Michigan, Ann Arbor, MI, USA; Department of Internal Medicine, Division of Gastroenterology, University of Michigan, Ann Arbor, MI, USA; Department of Neurosurgery, University of Michigan, Ann Arbor, MI, USA

## Abstract

The brain avidly consumes glucose to fuel neurophysiology. Cancers of the brain, such as glioblastoma (GBM), lose aspects of normal biology and gain the ability to proliferate and invade healthy tissue. How brain cancers rewire glucose utilization to fuel these processes is poorly understood. Here we perform infusions of ^13^C-labeled glucose into patients and mice with brain cancer to define the metabolic fates of glucose-derived carbon in tumor and cortex. By combining these measurements with quantitative metabolic flux analysis, we find that human cortex funnels glucose-derived carbons towards physiologic processes including TCA cycle oxidation and neurotransmitter synthesis. In contrast, brain cancers downregulate these physiologic processes, scavenge alternative carbon sources from the environment, and instead use glucose-derived carbons to produce molecules needed for proliferation and invasion. Targeting this metabolic rewiring in mice through dietary modulation selectively alters GBM metabolism and slows tumor growth.

**Significance:** This study is the first to directly measure biosynthetic flux in both glioma and cortical tissue in human brain cancer patients. Brain tumors rewire glucose carbon utilization away from oxidation and neurotransmitter production towards biosynthesis to fuel growth. Blocking these metabolic adaptations with dietary interventions slows brain cancer growth with minimal effects on cortical metabolism.

## Introduction

Gliomas are the most common form of malignant brain tumor, arising when normal glial cells of the central nervous system transform to become aggressive and invade the brain. Glioblastoma (GBM) is the most common aggressive type of brain cancer and characterized by profound invasiveness and treatment resistance. Conventional GBM treatment consists of surgical resection followed by radiation therapy (RT), temozolomide (TMZ) chemotherapy and sometimes tumor treating fields. Despite these treatments, GBMs invariably recur, and most patients die within 1-2 years of diagnosis^1–5^.

Poor outcomes for patients with GBM and other forms of glioma are due largely to treatment resistance, as the extensive inter- and intratumoral genomic heterogeneity of tumors limits therapeutic efficacy^6,7^. Further understanding of common targetable phenotypes in glioma might advance our efforts to develop novel therapeutics and improve the effectiveness of current standard treatments. Cancer cells exhibit dramatic differences in metabolic activity relative to neighboring healthy cells and rewire the flow of metabolism to favor proliferation and treatment resistance^8^. Moreover, we and others have shown that in glioma altered metabolism mediates a variety of important pro-tumor processes including treatment resistance^9,10^. Thus, targeting tumor metabolism for therapeutic benefit in patients is an attractive clinical strategy.

Defining metabolic pathway activity in human cancer is challenging. Positron emission tomography with glucose analogs shows high glucose uptake in both GBM and normal cortex^11^, but provides no information on how these tissues differentially utilize glucose-derived carbons. Quantifying metabolite abundance (such as through mass spectrometry-based metabolomics or magnetic resonance spectrometry) can reveal differences in metabolite levels between tumors and brain tissue. However, these steady state measurements provide minimal information regarding metabolic pathway activity, as the level of a metabolite is a function of both its rate of production and its rate of consumption. For example, high levels of a given metabolite could reflect its increased synthesis (increased activity) or its decreased consumption (decreased activity). Because these disparate biologic states are likely to respond differently to therapeutic inhibition, it is critical to understand the metabolic pathways that are active in a cancer to appropriately guide metabolically directed therapy.

Isotope tracing allows for the direct interrogation of metabolic pathway *activity* in cancer. In this technique, metabolic substrates containing heavy (but non-radioactive) isotopes such as ^13^C and ^2^H are administered to living systems. Tracking these isotopes into their downstream fates by mass spectrometry or nuclear magnetic resonance-based methods yields information about which metabolic pathways are active in a system. Applying this methodology to human cancers has revealed suppressed glucose oxidation in kidney cancer^12^ and enhanced lactate utilization, glycolysis and glucose oxidation in lung cancer^13,14^. Limited applications of this technology to human brain cancer have shown that the TCA cycle is active and can be fueled by both glucose and acetate^15,16^. How active other metabolic pathways are in human brain cancer and how brain cancer metabolism differs from that of cortex are unanswered questions.

To answer these questions, we infused ^13^C-labeled glucose into mice bearing orthotopic GBMs and patients with high-grade gliomas and followed the fates of glucose carbon into numerous downstream metabolic pathways in both cancerous and cortical brain tissue. We paired these mass spectrometry-based measurements with newly developed *in vivo* metabolic flux models to quantify the absolute rates of numerous metabolic reactions. Our studies reveal that aggressive brain cancers shift glucose carbon utilization away from physiologic processes such as TCA cycle oxidation and neurotransmitter synthesis, in part by salvaging nutrients like serine from the environment. Instead, they preferentially utilize glucose carbons to synthesize the molecules they need to grow: purines, pyrimidines, and nicotinamide cofactors. We also find that this adaptive metabolic regulation is plastic, with GBMs adaptively upregulating these metabolic pathways in response to therapy. Restricting alternative carbon sources by modulating diet shifts GBM metabolism away from biomass production and slows tumor growth. Together, these studies represent the first measurements of numerous metabolic pathways in human cancer and reveal brain cancer-specific metabolic rewiring that can be selectively targeted with dietary approaches.

## Results

### 13C glucose infusions into glioma patients and mouse models of GBM

To understand the metabolic fates of glucose in brain tumors, we infused uniformly labeled ^13^C-glucose ([U^13^C]-glucose) into mice with intracranial GBM patient-derived xenografts (PDXs) and into patients with likely GBM undergoing surgical resection. We then analyzed tissue by mass spectrometry to determine the accumulation of glucose-derived ^13^C into downstream metabolites (**Fig 1A**)^17^. In mice, we utilized a treatment-resistant luciferase^+^ GFP^+^ PDX (GBM38) from the Mayo Clinic repository^18^ and separated GFP^+^ tumor from GFP^-^ cortex using fluorescent-guided microdissection. In patients, sample isolation was performed using MRI guidance by a board-certified neurosurgeon (WNA). Our current surgical practice for glioma patients with tumors in non-eloquent locations is to perform a supramaximal resection which removes the contrast-enhancing tumor, the non-enhancing fluid-attenuated inversion recovery (FLAIR) hyperintense tumor, and some surrounding cortex (**Fig 1B**)^19^.

**Figure 1.**
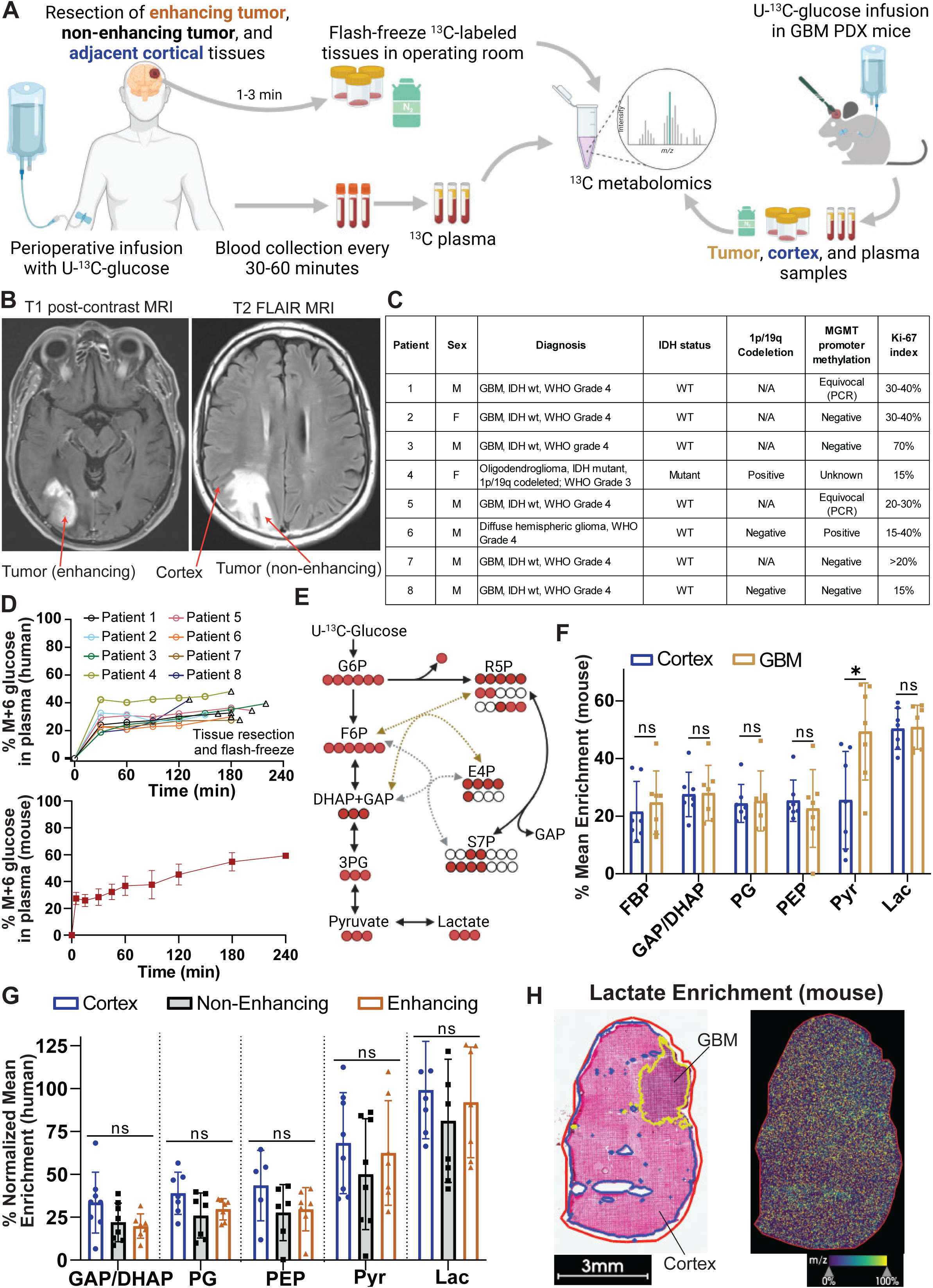
Robust [U^13^C]-glucose uptake and utilization in mouse models and patients with brain cancer. (A) Schema of [U^13^C]-glucose infusions into patients and mouse models of brain cancer. (B) Example of MRI-defined tissue acquisition. (C) Clinical and molecular characteristics of patients studied with stable isotope tracing. (D) Time course of M+6 arterial glucose enrichment in patients (top) and mice (bottom) undergoing [U^13^C]-glucose infusions. For mice, error bars indicate SDfrom 2-10 mice. (E) Schema of glucose carbon (red circles) redistribution into glycolytic intermediates. Scrambling can occur via recombination with unlabeled intermediates in the pentose phosphate cycle (E4P, S7P, R5P, GAP). (F) Mean enrichment of glycolytic intermediates in GBM (gold) and cortex (blue) tissue isolated from orthotopic GBM38-bearing mice infused with [U^13^C]-glucose (n=7 mice, error bars indicate SD). (G). Normalized (to plasma M+6 glucose on a per-patient basis) enrichment of glycolytic intermediates in cortex (blue), non-enhancing tumor (grey) and enhancing tumor (orange) from 8 patients infused with [U^13^C]-glucose (n=7-8 samples per group, error bars indicate SD). (H) H&E staining of GBM38 PDX grown orthotopically (left) and MALDI image showing ^13^C enrichment of lactate with tissue maximum set at 100%. * p<0.05. Metabolite abbreviations as follows: FBP (Fructose Bisphosphate), GAP (Glyceraldehyde phosphate), DHAP (dihydroxyacetone phosphate), PG (phosphoglycerate), PEP (phosphoenolpyruvate), Pyr (pyruvate), Lac (lactate), G6P (glucose 6-phosphate), F6P (Fructose 6-phosphate), R5P (ribose 5 phosphate), E4P (erythrose 4-phosphate), S7P (sedoheptulose 7-phosphate), 3PG (3-phosphoglycerate).

Eight patients enrolled in this study (**Fig 1C**), 6 of whom later were found to have GBMs, one of whom had an isocitrate dehydrogenase (IDH) mutant anaplastic oligodendroglioma, and one of whom had a histone H3 mutant G34R grade 4 glioma. Cortex and non-enhancing tumor were obtained from all 8 patients. Enhancing tumor was obtained from only 7 patients, because one tumor was comprised of entirely non-enhancing disease.

In human patients, [U^13^C]-glucose infusions lasted for the duration of the craniotomy, which was typically around 3 h. Circulating arterial [U^13^C]-glucose levels ranged between 20-40% of total arterial glucose during surgery and were typically at steady state after 30 minutes (**Fig 1D**). ^13^C labeling of arterial lactate (formed from tissue conversion of infused labeled glucose into lactate, which is then secreted into the circulation) varied between patients and was typically between 10-30% (**Fig S1A**). As in patient infusions, labeled glucose reached arterial steady state in mice within 30 minutes and a total label abundance of around 50% (**Fig 1D**), while ^13^C labeling of circulating lactate reached approximately 30% (**Fig S1B**).

We used hematoxylin and eosin (H&E) staining to confirm adequate separation of human samples (**Fig S1C**). Tumor content quantification by a board-certified neuropathologist (S.V.) indicated approximately 80% tumor content in enhancing tumor samples, 80% cortex in cortex samples and a mixture in non-enhancing tumor samples (**Fig S1D**). Consistent with these results, human cortex had nearly 10-fold higher levels of N-acetylaspartate (NAA) than enhancing tumor (**Fig S2A**), in linewith MR spectroscopy reports^20,21^ thereby further supporting adequate surgical separation of the tissues. In agreement with prior reports^22–24^, numerous other metabolites significantly differed in absolute levels between cortex and tumor tissue in both mouse and patients (**Fig S2B-D**).

### Glioma and cortex have similar glucose carbon incorporation into glycolytic intermediates

Following its entry into tissues, glucose is metabolized through glycolysis and the pentose phosphate cycle, allowing its carbons to be utilized for numerous downstream fates. Monitoring ^13^C carbon incorporation into metabolites of these pathways allowed us to determine pathways active in glioma and cortical tissues (**Fig 1E**). In tumor bearing mice (**Fig 1F**) and humans (**Fig 1G**), metabolites involved in upper glycolysis (fructose bisphosphate, GAP/DHAP, phosphoglycerate, PEP) displayed similar levels of ^13^C labeling in both GBM and cortex, suggesting that labeled glucose adequately and rapidly reached both tissues. Consistent with this finding, UDP-glucose, which is rapidly formed from glucose-1-phosphate and UTP, was similarly labeled in tumor and non-tumor tissues in mice and patients (**Fig S3A,B**). These findings are consistent with previous reports showing robust FDG uptake in cortex and GBM using PET imaging^25^.

Interestingly, in both GBM and cortex, the enrichment of lactate and pyruvate were higher than the enrichment of upstream glycolytic intermediates (**Fig 1F,G**). These labeling patterns resemble those seen in lung cancer^14^ and suggest possible ^13^C entry into lower glycolysis through lactate uptake or exchange.

To complement these LC-MS-based analyses, we also analyzed slices of flash-frozen tumor tissue by matrix-assisted laser desorption/ionization (MALDI) mass spectrometry. This technique has the advantage of analyzing metabolite levels in non-homogenized tissue samples and thus does not require cortex/tumor separation^26^. However, it lacks some of the sensitivity and metabolite-identification properties of LC-MS. Consistent with our LC-MS results, the glycolytic product lactate had similar ^13^C enrichment in GBM tissue and cortex (**Fig 1H** **and Fig S4A,B**). Together, these data indicate adequate ^13^C glucose entry into tumor and cortical tissues during infusions and similar utilization/exchange of labeled extracellular lactate.

### Gliomas reduce glucose-driven TCA cycle activity and neurotransmitter biosynthesis

Using these same samples, we then investigated metabolites in the TCA cycle, a central metabolic hub that allows for both the oxidation of glucose-derived carbons and their conversion into other molecules such as neurotransmitters and amino acids (**Fig 2A**). In mouse (**Fig 2B**) and human (**Fig 2D**) cortex, tracer-derived ^13^C accounted for approximately 30-40% of the TCA cycle intermediates citrate/isocitrate, α-ketoglutarate, succinate, and malate. By contrast, in tumor tissue, ^13^C accounted for only 15-20% of these TCA cycle metabolites (**Fig 2B,D**). This decrease in ^13^C labeling may indicate decreased TCA cycle activity in GBM compared to cortex and/or preferential utilization of non-glucose carbon sources to fuel the TCA cycle^27–32^.

**Figure 2.**
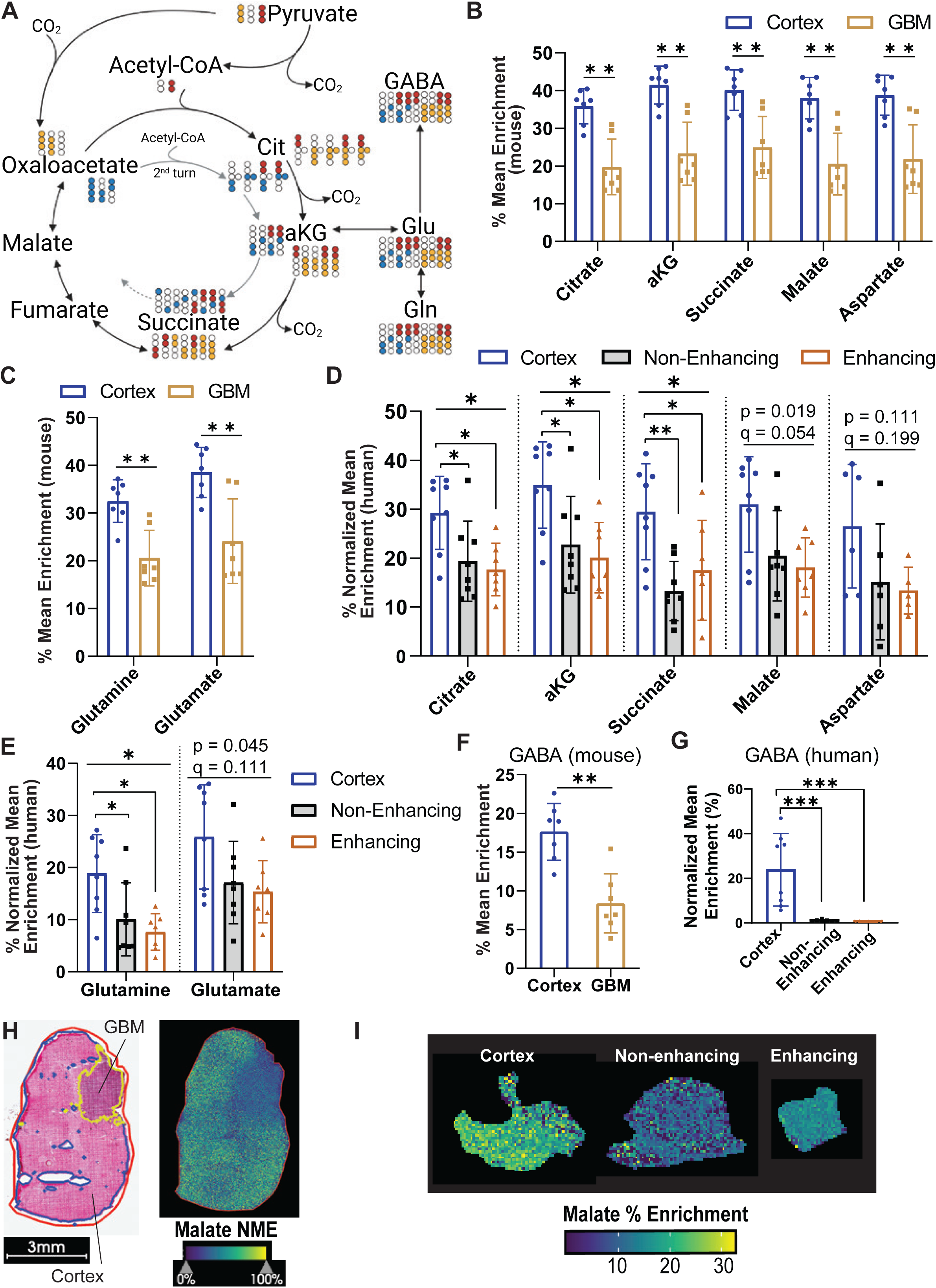
Decreased TCA cycle and neurotransmitter labeling in brain cancer. (A) Schema of ^13^C labeling in TCA cycle intermediates and neurotransmitters arising from M+3 pyruvate. Red circles indicate entry through pyruvate dehydrogenase, and orange indicates entry through pyruvate carboxylase. Blue circles indicate labeling patterns possible on second TCA cycle turn. (B) Mean enrichment of TCA cycle intermediates and aspartate in GBM (gold) and cortex (blue) tissue isolated from orthotopic GBM38-bearing mice infused with [U^13^C]_-_glucose (n=7 mice, error bars indicate SD). (C) Mean enrichment of glutamine and glutamate in GBM (gold) and cortex (blue) tissue isolated from orthotopic GBM38-bearing mice infused with [U^13^C]-glucose (n=7 mice, error bars indicate SD). (D) Normalized (to plasma M+6 glucose on a per-patient basis) enrichment of TCA cycle intermediates and aspartate in cortex (blue), non-enhancing tumor (grey) and enhancing tumor (orange) from 8 patients infused with [U^13^C]-glucose (n=7-8 samples per group and error bars indicate SD). (E) Normalized (to plasma M+6 glucose on a per-patient basis) enrichment of glutamate and glutamine in cortex (blue), non-enhancing tumor (grey) and enhancing tumor (orange) from 8 patients infused with [U^13^C]-glucose (n=7-8 samples per group, error bars indicate SD). Mean enrichment (F) and normalized mean enrichment (G) of GABA in mouse (F) and human (G) samples. (H) Malate normalized mean enrichment by MALDI. Color bar tissue maximum normalized to 100%. (G) Malate % enrichment in representative human cortex (left) non-enhancing tumor (middle) and enhancing tumor (right). Color bar maximum set at true enrichment. Abbreviations: aKG (alpha-ketoglutarate), NME (normalized mean enrichment). * p<0.05, ** p<0.01.

We then investigated neurotransmitter synthesis, which is critical for the functioning of normal brain and oncogenic phenotypes in brain cancer^33^. Glucose-derived carbons comprised a significant fraction of the neurotransmitters glutamate and gamma-aminobutyric acid (GABA) (**Fig 2C, E-G**). In both human patients and mouse models, tumor tissue had lower ^13^C labeling of glutamate, glutamine, and GABA than normal cortex, with GABA labeling in human GBM virtually absent (**Fig 2G**). Thus, we conclude that GBMs utilize less glucose to drive neurotransmitter synthesis than does normal cortex. These measurements were largely corroborated by our separate MALDI-based analysis which similarly revealed decreased ^13^C labeling in TCA cycle intermediates, glutamate, and glutamine (**Fig 2H-I, Fig S4C-F, Fig S5A-C, Fig S6A-C**).

### Gliomas increase glucose carbon incorporation into nucleotides

With similar glucose uptake yet lower glucose contributions to the TCA cycle and neurotransmitter synthesis, we next sought to determine how GBMs preferentially utilized glucose-derived carbons. We interrogated the labeling patterns of the glucose-derived metabolites that cells use as building blocks to synthesize the macromolecules they need to grow and divide.

Nucleotides and their derivatives are comprised of purines and pyrimidines, which are produced through separate metabolic pathways that both use glucose-derived ribose 5-phosphate (R5P) produced in the pentose phosphate pathway (**Fig 1E**). Cells may generate purines *de novo*, in which carbon and nitrogen from numerous sources are added to R5P in an energy-intensive process (**Fig 3A**). Alternatively, cells can also salvage nucleotides by conjugating R5P with pre-formed nucleobases. (**Fig 3A**). Notably, ^13^C labeling of many purine metabolites, including GMP and GDP, was increased in brain cancer relative to cortex in both mouse models (**Fig 3B**) and human patients (**Fig 3C**). Since the enrichment of R5P is similar between the tissues, the increase in purine ^13^C enrichment is most likely a result of higher synthesis in gliomas (**Fig S7A-B**). We observed similar results using MALDI, which specifically showed increased ^13^C-labeling of purines in GBM tissue compared to normal cortex (**Fig 3H, S4G)**. Interestingly, when we evaluated the enrichment patterns of individual patients, we found that only the GMP arm of the purine pathway had consistently higher enrichment in all patients (**Fig S8A-D, S9A-C**). This indicates that GMP production might be especially important for gliomas.

**Figure 3.**
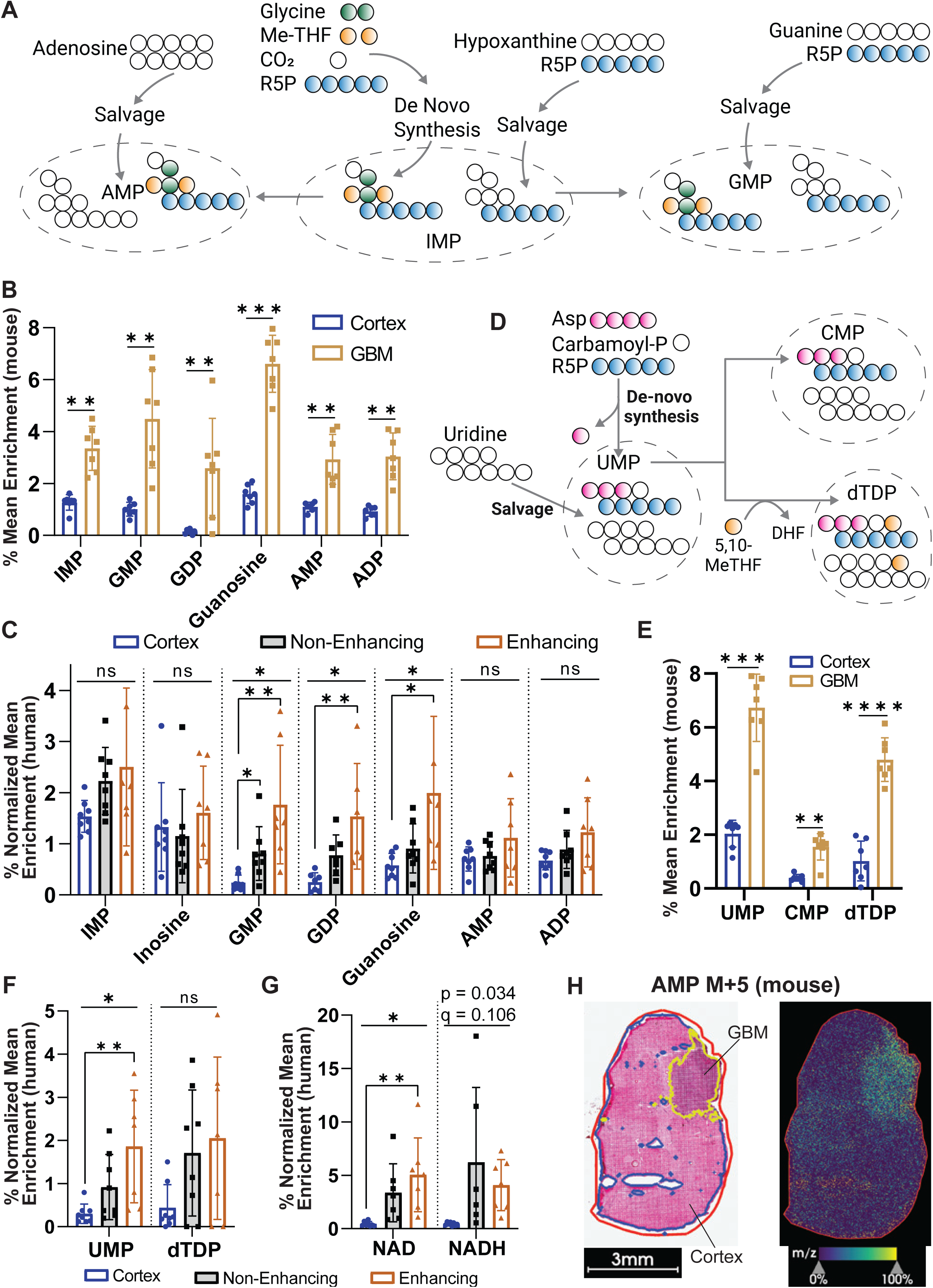
Increased glucose-driven nucleotide synthesis in brain cancer. (A) Schema of purine synthetic pathways. Green and yellow circles indicate glycine- and folate-derived carbons, respectively. Blue circles indicate ribose 5-phosphate (R5P)-derived carbons. Partial shading of these circles indicates that a variety of labeling patterns are possible. (B) Mean enrichment of purines in GBM (gold) and cortex (blue) tissue isolated from orthotopic GBM38-bearing mice infused with [U^13^C]-glucose (n=7 mice, error bars indicate SD). (C) Normalized (to plasma M+6 glucose on a per-patient basis) enrichment of purines in cortex (blue), non-enhancing tumor (grey) and enhancing tumor (orange) from 8 patients infused with [U^13^C]-glucose (n=7-8 samples per group, error bars indicate SD). (D) Schema of pyrimidine synthetic pathways. Purple indicates aspartate-derived carbons, blue indicates R5P-derived carbons, and yellow indicates folate-derived carbons. Partial shading is used to indicate that a variety of labeling patterns are possible (E) Mean enrichment of pyrimidines in GBM (gold) and cortex (blue) tissue isolated from orthotopic GBM38-bearing mice infused with [U^13^C_6_]_-_ glucose (n=7 mice, error bars indicate SD). (F) Normalized (to plasma M+6 glucose on a per-patient basis) enrichment of pyrimidines in cortex (blue), non-enhancing tumor (grey) and enhancing tumor (orange) from 8 patients infused with [U^13^C]-glucose (n=7-8 samples per group, error bars indicate SD). (G) Normalized (to plasma M+6 glucose on a per-patient basis) enrichment of NAD and NADH in cortex (blue), non-enhancing tumor (grey) and enhancing tumor (orange) from 8 patients infused with [U^13^C]-glucose (n=7-8 samples per group, error bars indicate SD). (H) AMP M+5 signal intensity by MALDI in mouse GBM and cortex. Color bar tissue maximum normalized to 100%. Abbreviations: Me-THF (N10-formyltetrahydrofolate). AMP (adenosine monophosphate), IMP (inosine monophosphate), GMP (guanosine monophosphate), GDP (guanosine diphosphate), ADP (adenosine diphosphate), 5,10-MeTHF (5,10-methylenetetrahydrofolate), UMP (uridine monophosphate), CMP (cytidine monophosphate), dTDP (deoxythymidine diphosphate), NAD (oxidized nicotinamide adenine dinucleotide), NADH (reduced nicotinamide adenine dinucleotide). * p<0.05, ** p<0.01, *** p<0.001.

Distinct from purine synthesis, pyrimidine production is initiated by formation of a nucleobase from aspartate and carbamoyl phosphate and then conjugated to R5P to eventually yield UMP (**Fig 3D**). UMP may alternatively be salvaged from uridine. UMP can be further metabolized to produce additional pyrimidines. Like purines, we found that pyrimidines have elevated ^13^C labeling in cancerous tissues compared to cortex in both mouse models (**Fig 3E**) and human patients (**Fig 3F**).

In addition to driving nucleotide synthesis, glucose-derived carbons are also used to form the essential cofactors NAD and NADH, both of which can promote oncogenic phenotypes (**Fig S11A**). Consistent with our findings of increased nucleotide labeling in glioma tissues compared to cortex, we additionally observed elevated labeling of both NAD and NADH in both mouse **(Fig S11B)** and human glioma (**Fig 3G**), indicating increased NAD/NADH synthesis that may support the increased growth and survival demands of tumors. Altogether these data show that gliomas rewire glucose carbon utilization away from TCA cycle oxidation and neurotransmitter synthesis and redirect them to fuel the biosynthetic needs of cancer growth.

### Gliomas increase flux of nucleotide synthesis and decrease oxidative TCA cycle flux

High ^13^C enrichment at a single time-point does not necessarily imply increased biosynthetic flux. For example, a nucleotide formed only from infused ^13^C-containing sources at a low rate might have higher ^13^C enrichment than a nucleotide formed from unlabeled sources at a faster rate. Therefore, we developed a metabolic flux analysis (MFA) approach to directly quantify nucleotide synthesis fluxes from *in vivo* enrichment data (**Fig 4A**)^34^. We then applied this mathematical framework to our patient-derived orthotopic GBM mouse models to determine if higher GBM nucleotide enrichment corresponds to higher synthetic flux.

**Figure 4.**
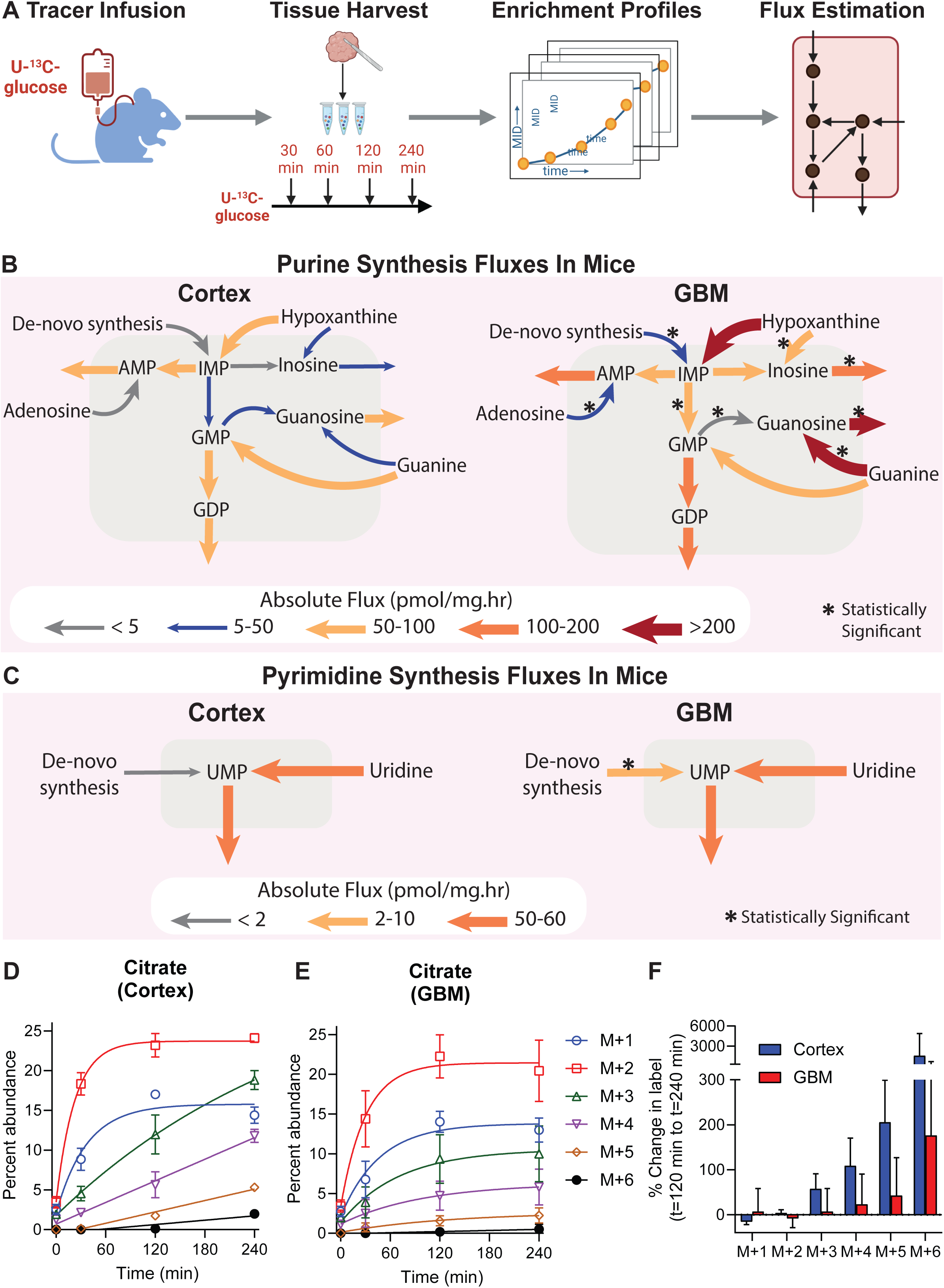
Quantification of metabolic fluxes in GBM and cortex. (A) Schema. [U^13^C]-glucose was infused into GBM38 PDX-bearing mice. Mice were serially euthanized at varying timepoints (0, 30, 120 and 240 min), and GBM and cortex was harvested to measure metabolite enrichment and estimate metabolic fluxes. (B-C) Absolute fluxes of purine (B) and pyrimidine (C) synthetic reactions in cortex (left) and GBM (right). * indicates p<0.05 comparing GBM and cortex. (D-E) Percent abundance of individual malate isotopologues in cortex (D) or GBM (E) Error bars indicate SD of n=3-9 datapoints from n=1-3 mice at each timepoint. (F) Percent change in indicated malate isotopologues from 120 to 240 min. Error bars are propagated from uncertainty in 120 and 240 min datapoints in (D) and (E).

We infused orthotopic GBM-bearing mice with [U^13^C]-glucose and harvested GBM and normal cortical tissue for LC-MS analysis at multiple time points post-infusion to generate time-dependent enrichment profiles for MFA (**Fig 4A, S12**). The time-course enrichment profiles of purine and pyrimidine nucleotides showed an increasing trend during entirety of the 4 h experiment (**Fig S12A-B**), suggesting that isotopic steady state in these pathways had yet to be achieved at time of tissue harvest in our single timepoint studies (**Fig 1-3**). To overcome this limitation, we applied an ordinary differential equation (ODE)-based MFA model using time-course nucleotide mass isotopomer distribution (MID) profiles to solve for fluxes (**Fig 4A**)^35^. We quantified 15 reactions in the purine synthesis pathway and 3 reactions in the pyrimidine synthesis pathway in GBM and adjacent normal cortex tissue (**Fig S13A-B, Tables S1-S4**).

Using MFA, we found significant differences in nucleotide synthesis rates between GBM and cortex. In the purine synthetic pathway, GBMs have higher *de novo* IMP and GMP synthesis fluxes than cortex, accompanied by increased salvage synthesis of IMP and AMP (**Fig 4B**, **Tables S9-S10**). We also noted increased synthesis of inosine, guanosine, and GDP in GBM, further highlighting the broad increase in purine biosynthesis in tumors. Notably, comparison of total metabolite concentrations in GBM and cortical tissues showed that many purines were present in equal or lower abundance in GBM compared to cortex (**Fig S12C**), further highlighting the increased information that can be obtained from ^13^C-MFA compared to metabolite isotope tracing and static metabolite levels-based analysis. Collectively these results indicate that the higher ^13^C enrichment of purines observed in tumors stems from higher absolute rates of purine synthesis.

Pyrimidine fluxes also differed between GBM and cortex. The *de novo* synthesis of UMP was elevated about 5-fold in GBM compared to cortex (**Fig 4C** **and Tables S11-S12**). However, uridine salvage was the dominant form of pyrimidine synthesis in both GBM and cortical tissue and accounted for more than 80% of UMP synthesis in both.

We also utilized our murine time-course studies to analyze TCA cycle activity in GBM and cortex. Entry of fully ^13^C_6_-labeled glucose-derived pyruvate into the TCA cycle through pyruvate dehydrogenase forms TCA cycle intermediates that have two ^13^C atoms (M+2, **Fig 2A**). These intermediates can then be oxidized through further turns in the TCA cycle, or they can exit the TCA cycle to drive other metabolic reactions. Intermediates that have experienced multiple turns of the TCA cycle can contain additional ^13^C atoms (M+3 to M+6, with M+3 also possible from a single turn involving the pyruvate carboxylase reaction, **Fig 2A**). In both GBM and cortex, we observed the rapid formation of M+2 TCA cycle intermediates (citrate, succinate and malate) over time. However, the abundance of M+3 to M+6 TCA cycle intermediates steadily rose over time in cortex but remained low in GBM (**Fig 4D-E, S14A-B, D-E**). Further, analyses of isotopologue distribution changes at the latest time points in our studies (t=120 min and t=240 min) in which GBM labeling approached steady state similarly revealed a consistent trend of higher labeling in cortex than GBM (**Fig 4F** **and Fig S14C,F**). These findings suggest higher oxidative turning of the TCA cycle in cortex and suggest that glucose-derived TCA cycle intermediates undergo less oxidation in GBM.

### GBM metabolic activity is dynamic following therapy

GBMs are characterized by profound treatment resistance. We therefore asked if their metabolic adaptations facilitated the ability to respond to standard of care treatments such as RT. We treated GBM-bearing mice with cranial RT delivered to the entire brain (tumor and cortex) immediately (<5 min) prior to [U^13^C]-glucose infusion and harvested tumor and cortex over time as in **Fig 4**.

Our initial MFA models (**Fig 4**) assume metabolic steady state throughout the infusion. This assumption is unlikely to be true after RT due to the rapid activation of the DNA damage response and subsequent cell cycle arrest. Therefore, we developed a dynamic-^13^C-MFA (DMFA) model to quantify purine synthesis fluxes after RT (**Fig 5A**).

**Figure 5.**
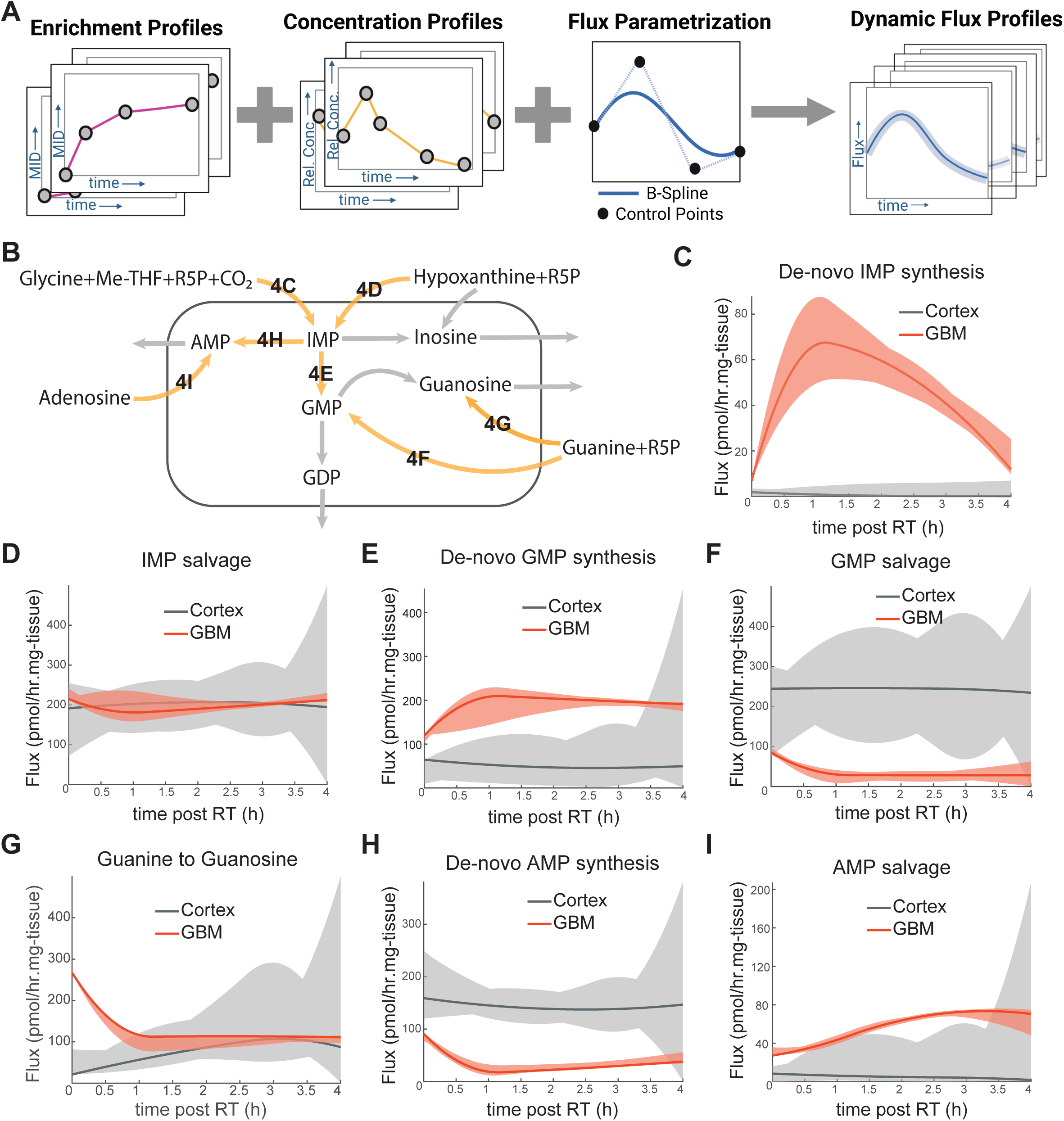
Dynamic metabolic response to radiation in GBM and cortex. (A) Schema of dynamic metabolic flux analysis pipeline. (B) Overview of purine synthesis pathways. (C-I) dynamic metabolic fluxes following RT in GBM (red) and cortex (grey). GBM38 PDX-bearing mice were treated with cranial RT (8Gy) at t=0. Mice were serially euthanized at varying timepoints after RT (0, 30, 60, 120 and 240 min), and GBM and cortex were harvested to estimate metabolic fluxes (n=1-3 mice per group with 1-3 samples/mouse). Solid lines indicate estimated fluxes and shaded regions indicate 95% confidence intervals. Abbreviations: Me-THF (again, I think this is *N*^10^ formyl-tetrahydrofolate), R5P (ribose 5-phosphate), IMP (inosine monophosphate), AMP (adenosine monophosphate).

A DMFA model incorporates time-dependent concentration changes and is used to estimate transient flux profiles^36,37^. We modified our *in vivo* ^13^C-MFA framework to include dynamic concentration and flux changes (**Fig 5A**, **S15A-B, Supplemental Methods**). Strikingly, purine fluxes changed dynamically after RT in GBM but remained largely unaffected in cortical tissue (**Fig 5B-I**, **Fig S16**). *De novo* IMP synthesis increased transiently after RT, with peak activity at approximately 1 h post-RT and diminishing over the next 3 h (**Fig 5C**). Notably, this pattern and timeframe of increased *de novo* IMP synthesis is well-aligned with the known timeframe of DNA damage and repair following RT^38^. In contrast, IMP salvage from hypoxanthine was unaffected in both GBM and cortex (**Fig 5D**). *De novo* synthesis of GMP from IMP also increased over approximately 1h, consistent with increased IMP synthesis, and this increase was sustained for the next 3h (**Fig 5E**), while guanylate salvage and *de novo* AMP synthesis both decreased after RT (**Fig 5F-H**). Increase in AMP salvage partially compensates for lower *de novo* AMP synthesis (**Fig 5I**). Together these data suggest that after RT, GBMs acutely increase *de novo* IMP synthesis that feeds into increased guanylate production with an accompanying decrease in AMP production.

### Gliomas preferentially rely on environmental serine

Synthesis of purines via the *de novo* pathway requires a variety of amino acid substrates including serine, which is a neurotransmitter, a driver of lipid synthesis and a precursor for glycine and one-carbon units. Because serine metabolism is important for a variety of tumors including GBM ^39–44^, we investigated whether our isotope infusions could help us understand serine metabolism in human brain cancer.

In both murine (**Fig 6A**) and human (**Fig 6B**) studies, total ^13^C labeling of serine was similar in brain cancer and cortex samples. This finding was distinct from many other amino acids, neurotransmitters and nucleotides, which exhibited differential label enrichment in brain cancer and cortex (**Fig 2,3**). Upon deeper investigation of labeling patterns in murine GBM and cortical tissue, we found that M+3 labeling of serine predominated in cortex while M+1 labeling predominated in GBM tissue (**Fig 6C**). We observed similar labeling patterns in most human tissues, in which M+3 serine was higher in cortex than enhancing tumor in nearly every case, while M+1 serine predominated in both enhancing and non-enhancing tumor tissue in most patients (**Fig 6D, S18A**). The predominance of M+1 serine has been seen in prior human cancer ^13^C_6_ glucose infusion studies but is of uncertain significance (**Fig S17A-E**). While cortex has not been analyzed in human isotope tracing studies, in mouse studies, the brain is the only organ in which M+3 labeling of serine predominates over M+1 in animals infused with ^13^C_6_ glucose (**Fig S17F**). These results suggest that our labeling patterns (increased M+1 serine in brain cancer, increased M+3 serine in cortex) are not spurious.

**Figure 6.**
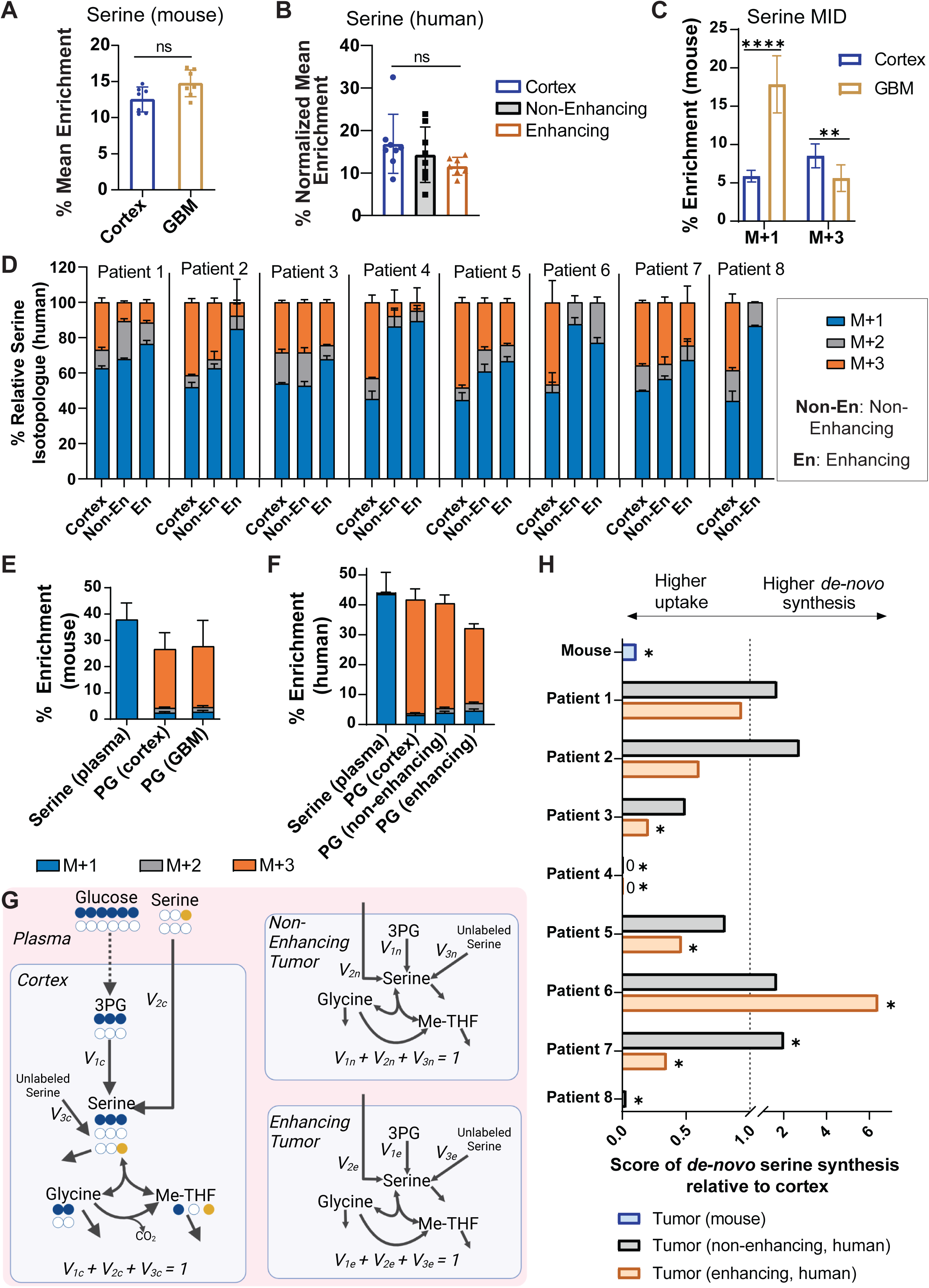
Preferential reliance on environmental serine in brain cancer. (A) Mean ^13^C enrichment of serine in cortex (n=7) or orthotopic GBM38 PDXs (n=7). (B). Normalized (to plasma glucose enrichment) mean enrichment of ^13^C in serine in human samples (n=7-8 per group). (C) Percent serine that contains one (M+1) or three (M+3) tracer-derived ^13^C atoms. (D) Serine isotopologue distributions of tissues from 8 human brain cancer patients. Error bars indicate SD of 3 technical replicates per patient. (E) Isotopologue distributions of plasma serine and tissue phosphoglycerate (PG) in PDXs. (F) Isotopologue distributions of plasma serine and tissue PG in human tissues (PG normalized to plasma glucose enrichment). (G) Metabolic flux analysis model to estimate routes of serine synthesis in cortex and brain cancer. (H) Relative (to cortex) reliance on serine uptake compared to glucose-driven *de novo* serine synthesis. Significance was tested by comparing the 95% confidence intervals from the flux model. * indicates p<0.05 compared to 1.0 (i.e., significantly different than cortex).

Labeled serine has multiple potential sources including (1) *de novo* synthesis from glucose, (2) uptake from the environment, and (3) synthesis from the addition of a carbon from the folate cycle with the two-carbon amino acid glycine. In both cortex and GBM tissue in mice, the glycolytic intermediate phosphoglycerate (PG), which is the precursor of serine, is almost exclusively M+3 labelled in both cortex and GBM (**Fig 6E**). Like in mice, human PG is predominantly M+3 labelled in all tissues (**Fig 6F**). Thus, any serine formed *de novo* from glycolytic intermediates would also be predominantly M+3 labelled. The predominance of M+3 serine in cortex and M+1 serine in brain cancer suggested that cortex predominantly generates its serine from glucose while GBM tissue likely relied on other sources.

We then investigated potential alternative serine sources for brain cancer. Arterial serine in both patients and mice was predominantly M+1 labelled (**Fig 6E, F**), likely due to synthesis from unlabelled glycine and a labelled folate carbon from the kidney and liver. Thus, higher reliance on uptake of circulating serine could give rise to the M+1 serine seen in murine and human brain cancer samples.

However, another potential source of M+1 is reverse SHMT flux, which can produce M+1 serine from the combination of unlabelled glycine and labelled 1-C units in the folate cycle (**Fig 6G**). Given this complexity, we developed multicompartment ^13^C-MFA model to understand the relative magnitudes of these fluxes in the patient tumors (**Fig 6H**). Our model was comprised of multiple tissue compartments, with all compartments having the same source of circulating serine (see methods). To ensure uniformity in the results, we constrained the external serine synthesis/uptake in all the compartments to 1. The resulting score of *de novo* serine synthesis flux to serine uptake flux is depicted in **Fig 6H** (calculation of the score is described in methods). A ratio higher than 1 would signify a higher contribution of *de novo* serine synthesis in the tumor relative to the cortex, while a ratio lower than 1 would mean higher reliance on serine uptake relative to the cortex. In mouse samples, cortex predominantly relied on *de novo* serine synthesis while GBM samples derived most serine from extracellular sources (**Fig 6H**). There was some heterogeneity in patient samples. While cortex predominantly generated serine *de novo*, many enhancing (6 of 7) and non-enhancing (4 of 8) tumor samples primarily relied on extracellular serine uptake (**Fig 6H, S18B**). Together, these data indicated that the relative flux of *de novo* serine synthesis is lower in many brain cancers compared to the cortex, and that brain cancers scavenge serine from the environment.

This observation suggested that brain cancers shift towards serine uptake and downregulate glucose-driven serine synthesis so that they can instead utilize glucose carbon for biosynthesis and growth. To test this hypothesis, we restricted environmental serine in orthotopic GFP^+^ fluc^+^ GBM-bearing mice by feeding them a serine-restricted diet (**Fig 7**). Dietary serine restriction decreased circulating serine levels as expected (**Fig S19A**) and significantly slowed tumor growth as assessed by bioluminescence (**Fig 7A,B**). When control mice neared a humane endpoint, we euthanized mice and harvested brain and tumor tissues from all groups for analysis. Tumors in serine-restricted mice were smaller than controls (**Fig 7C**) and had a lower proliferation index as measured by Ki-67 staining (**Fig 7D-E**). Metabolite quantification of showed that the serine/glycine restricted diet dramatically altered the metabolome of GBM tissues as assessed by principal component analysis but had little effect on cortical metabolism (**Fig S19B, S19C)**. This observation is consistent with glucose-driven *de novo* serine synthesis being the predominant route of synthesis in cortex. GBM samples from mice on low serine/glycine diets had lower nucleotides, NAD^+^ and NADH compared to GBM samples (**Fig 7F**) but only modestly decreased serine levels (**Fig 7G**). These findings suggested a compensatory re-routing of glucose carbons towards serine within the tumor when extracellular serine sources were limiting. Consistent with this hypothesis, phosphoserine levels (an intermediate formed when glucose carbons are shunted towards serine) were dramatically elevated in GBM tissue from mice fed a serine/glycine restricted diet (**Fig 7G**). Together these data indicate that many brain cancers preferentially rely on extracellular sources for serine but can adapt to serine restriction by slowing proliferation and rerouting glucose carbons away from biomass production and towards serine synthesis (**Fig 7H-J**).

**Figure 7.**
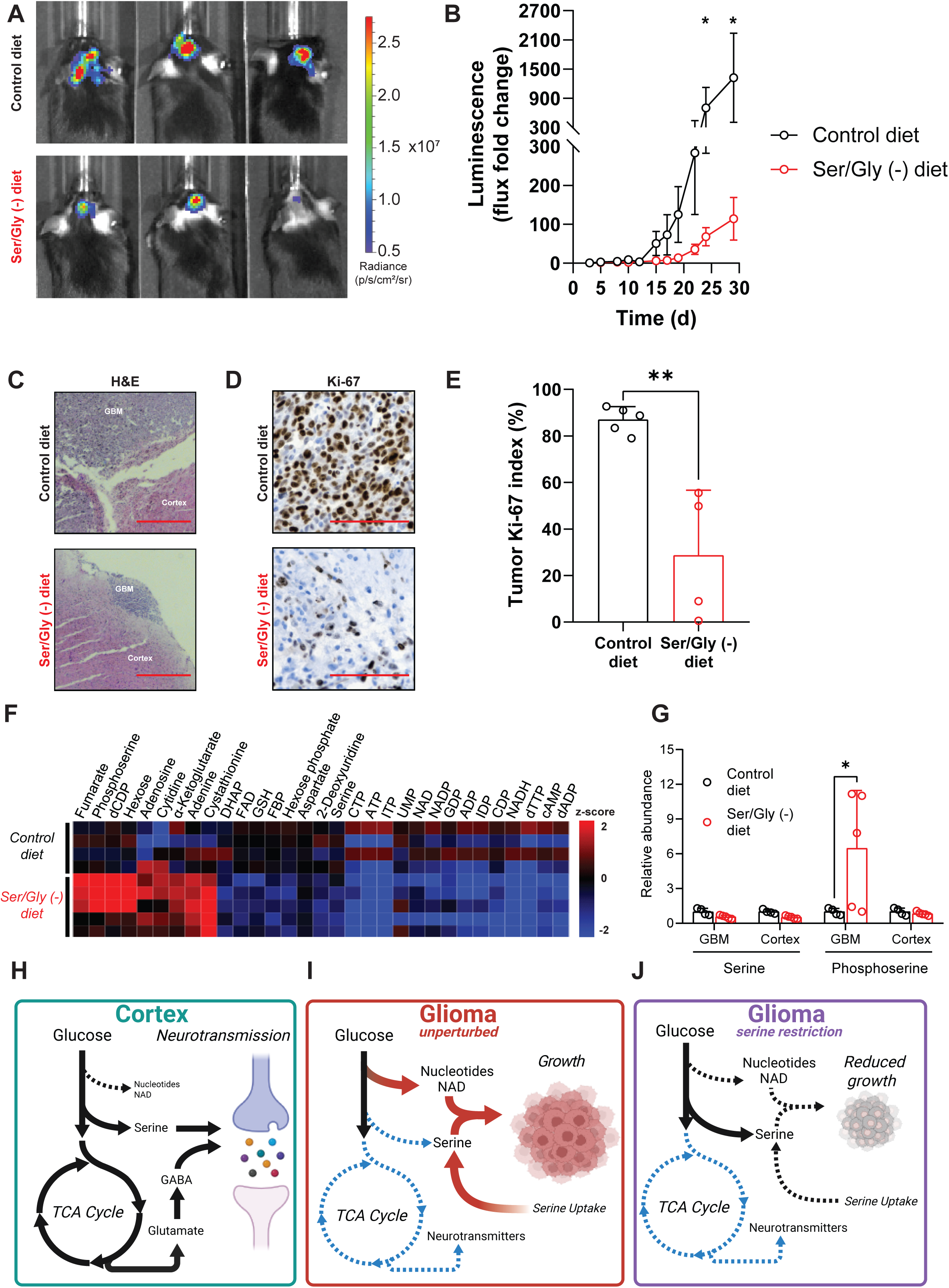
Dietary serine restriction selectively alters GBM metabolism and slows tumor growth. (A) Representative bioluminescence imaging of GBM38 PDXs grown orthotopically in mice on control (top) or serine/glycine (Ser/Gly) -restricted diet (bottom). (B). Fold-change in luminescence (compared to mean luminescence on day 3) in GBM38 PDX-bearing mice fed a control diet (black) or Ser/Gly-restricted diet (red). Error bars indicate SEM from 9-10 animals per group. (C-G) Mice in panel B were euthanized approximately 4 weeks after implantation, and brains were bisected for analysis by histopathology (C-E) or metabolomics (F-G). (C-D). Representative H&E (C) or Ki-67 immunohistochemistry (D) images from mice on control or Ser/Gly restricted diets. (E) Ki-67 quantification of GBM38 tumors from mice on a control diet (black, n=5) or Ser/Gly-restricted diet (red, n=4). (F) Heatmap of relative metabolite levels (top 65 PLS-DA) in orthotopic GBM38 tumors from mice fed a control (n=4) or Ser/Gly-restricted diet (n=5), with each row representing a separate tumor. (G) Relative serine and phosphoserine levels in GBM38 tumors and cortex from mice fed control or Ser/Gly-restricted diets. Error bars indicate SD. (H-J) Models of cortical metabolic rewiring in brain cancer. (H) Cortex robustly takes up glucose, which it uses to fuel the TCA cycle and the synthesis of neurotransmitters serine, GABA, and glutamate. (I) Brain cancers upregulate the uptake of environmental serine and downregulate the oxidation of glucose in the TCA cycle and glucose-derived neurotransmitter synthesis. Tumors also re-route glucose-derived carbons to synthesize nucleotides and NAD/NADH, used to drive tumor growth. (J) Restriction of dietary serine forces gliomas to re-route glucose carbon towards serine synthesis, which decreases nucleotide and NAD/NADH levels and slows tumor growth. * p<0.05, ** p<0.01.

## Discussion

In this work, we establish a clinical stable isotope tracing program which integrated ^13^C based infusions with ^13^C-MFA to define the metabolic rewiring that occurs in brain cancer and understand its therapeutic implications. While both brain cancer and cortex avidly consume glucose and engage in glycolysis, cortex predominantly utilizes glucose-derived carbons for physiologic processes such as TCA cycle oxidation and neurotransmitter synthesis. Brain cancers downregulate these physiologic processes and instead use glucose-derived carbons to synthesize nucleotides including NAD^+^ and NADH, which they use to fuel proliferation. By developing quantitative ^13^C-MFA models, we identify upregulated *de novo* purine and pyrimidine synthesis in GBM and a robust GBM-specific metabolic response to radiation therapy. Finally, we reveal that cortex synthesizes a higher fraction of its serine from glucose, while brain cancers salvage serine from the environment. This metabolic rewiring is a targetable liability with a therapeutic window. In mouse models, dietary serine restriction depletes GBM nucleotide pools and slows tumor growth while minimally affecting the metabolism of the cortex.

Our work provides insights into how metabolism is altered in human brain cancers and adds to a growing body of knowledge regarding metabolic rewiring in cancer. Like the radioactive glucose analogs used in positron-emission tomography, we find that infused [U^13^C]-glucose rapidly accumulates in both GBM and cortex^25^. Our findings suggest that broadly targeting glucose uptake in attempt to slow GBM growth is unlikely to have a favorable therapeutic window and may lead to untoward toxicity. Like previous studies with ^13^C-labeled nutrients, we find an active TCA cycle in brain cancer^15,16^. However, our unique surgical practice allowed us to perform the first comparisons of metabolic pathway activity in human cortex and brain cancer. While the TCA cycle is active in GBM, it appears downregulated compared to non-malignant cortex with both a preference for non-glucose substrates and decreased turning of the cycle. To our knowledge, routes of serine synthesis have not been interrogated in human cancer. Unlike brain metastases, which appear to rely on *de novo* serine synthesis in preclinical models^43^, GBMs preferentially rely on environmental serine, and do so to allow glucose carbon to be used for nucleotide synthesis and proliferation instead.

The ^13^C-MFA models we develop in this work provide insights into metabolic rewiring that are difficult to glean with simpler analysis techniques^34,35,45–47^. While *de novo* synthesis of both purines and pyrimidines is elevated in GBM compared to cortex, salvage of uridine and hypoxanthine appears to dominate nucleotide production. This situation appears to differ from that of other brain tumors, where the *de novo* synthesis of pyrimidines is critical^48–50^. These results suggest that targeting the upstream steps of *de novo* nucleotide synthesis in GBM may lack efficacy due to compensatory salvage pathways that can fill nucleotide pools when upstream steps are blocked. Our flux models also indicate that GBMs adaptively rewire metabolism in response to radiation, perhaps explaining why these tumors typically recur following radiation treatment. Further, the dynamic ^13^C-MFA analysis revealed an increased reliance on *de novo* GMP synthesis after radiation, highlighting the important role that metabolic fluxes can play in treatment responses. Some of our flux models require sample acquisition at multiple timepoints and are not suitable for clinical application where tumor is typically removed only once. However, our serine production flux model requires only a single timepoint, so we are hopeful that it and similar models will have increased applicability in real world practice. Our current serine production model makes several simplifying assumptions (described in supplementary methods), as our ability to add complexity to the model depends on the underlying data. Our study demonstrates the utility of simplified models in extracting quantitative information from metabolic studies. As experimental techniques grow, the data can be used to develop more complex and representative models.

Our work has important clinical implications. Inhibiting nucleotide synthesis, serine uptake, or non-glucose TCA cycle fuel sources might have a therapeutic index to selectively affect GBM, whereas broadly targeting glucose uptake may cause unacceptably cortical toxicity. Targeting proximal *de novo* nucleotide synthesis in GBM may be ineffective due to active salvage pathways. Blocking IMPDH may still have therapeutic benefit in glioma with a favorable therapeutic ratio because of the preference for gliomas to salvage hypoxanthine. Restricting dietary serine could help slow GBM growth and potentially augment the efficacy of standard of care GBM treatments^51^, though the efficacy of this approach could be limited by local production of serine in the GBM microenvironment^52^.The patient-to-patient heterogeneity in environmental serine dependence we observed suggests that isotope tracing could be used a precision medicine technique to determine which patients are mostly likely to benefit from dietary serine restriction.

Our study lays the groundwork for future investigation. Because we performed isotope tracing on only 8 patients, we are not yet able to correlate individual mutations to metabolic activity. Increasing the utilization of isotope tracing beyond specialized centers and into wider clinical practice will help us understand links between genotype and metabolic phenotype, understand causes of metabolic heterogeneity, and understand whether there is prognostic and predictive value in isotope tracing data. Using tracers beyond glucose will help us understand additional aberrant metabolic pathways in cancer. And as spatial metabolomic techniques advance and reach single cell resolution, we may be able to spatially resolve these metabolic fluxes to understand metabolic dysregulation in discrete cell types in the GBM microenvironment. Further, by overlaying phenotypic data from spatial transcript or protein analysis, we may better understand how metabolism relates to growth, cell state and treatment resistance.

In summary, we have discovered a profound rewiring of carbon metabolism in aggressive human brain cancers that fuels tumor growth. These cancer-specific metabolic alterations, such as the preference for environmental serine and the reliance on IMPDH to synthesize GTP, are potential therapeutic targets with favorable therapeutic indices. Our research team and others are currently inhibiting these metabolic pathways in patients with brain cancer (NCT04477200, NCT05236036). We are hopeful that the growing use of isotope tracing in patients with cancer will help us make additional fundamental observations about cancer metabolism and teach us which patients might benefit most from which metabolically targeted therapy.

## Methods

### Clinical stable isotope tracing protocol

Eight patients with suspected GBM were enrolled onto our IRB-approved clinical study, which was performed perioperatively with standard-of-care craniotomies for tumor resection. Near the start of each procedure (approximately 2-4 hours prior to tissue resection), patients received a bolus intravenous dose of [U^13^C]-glucose (8 g) followed by a continuous intravenous infusion of [U^13^C]-glucose at a rate of 4 g/h. Arterial blood was collected into EDTA-coated vials every 30-60 min for plasma preparation and analysis until solid tissues of interest were harvested from each patient. After initial craniotomy and tumor exposure, stereotactic image guidance was utilized to identify radiographically defined enhancing tumor, non-enhancing FLAIR T2 hyperintense tumor, and adjacent cortical tissues. These tissues were then resected and separated by the neurosurgeon (WNA), rinsed in cold PBS and immediately (<3 min after resection) flash-frozen in liquid nitrogen by the research team for further analysis.

### Animal studies and patient-derived xenograft model

All animal studies were approved by the University Committee on Use and Care of Animals at the University of Michigan. In all animal experiments, male and female mice aged 4-12 weeks were used. Mice were housed in specific pathogen-free conditions at a temperature of 74 °F and relative humidity between 30 and 70% on a light/dark cycle of 12 h on/12 h off with unfettered access to food (PicoLab® Laboratory Rodent Diet, 5L0D) and water.

Studies assessing intracranial tumor-bearing mice used the PDX model GBM38, which is a chemoradiation resistant model that genetically and histologically represents a typical GBM, from the PDX National Resource at Mayo Clinic^53^. Tumor tissue was propagated subcutaneously in the flanks of immunodeficient mice (B6.129S7-Rag1^tm1Mom^/J [Rag1 KO], Jackson Laboratory). To introduce GFP and fluc into GBM tissue, flank tumors were used to generate short-term explant cultures and transduced by lentiviral infection (lenti-LEGO-Ig2-fluc-IRES-GFP-VSVG). Following infection, cells were enriched for GFP^+^ populations by fluorescence-activated cell sorting and then reintroduced to mice either as subcutaneous flank tumors or intracranial tumors. To generate orthotopic GBM brain tumors, 5×10^5^ GFP^+^ fluc^+^ GBM38 cells were stereotactically implanted into the region of the brain calculated to be the striatum in anesthetized Rag1 KO mice. Tumor development was then confirmed by bioluminescence imaging (BLI) as described below.

### Tumor growth and bioluminescence

To monitor intracranial tumor growth in mice, we employed BLI, which leverages the expression of luciferase in intracranial tumors. For each measurement, mice intracranially implanted with fluc^+^ GBM38 cells were intraperitoneally injected with 150 mg/kg D-luciferin. Ten minutes after injection, mice were imaged using an IVIS™ Spectrum imaging system (PerkinElmer) while under anesthesia (2% isoflurane inhalation). In tumor growth studies, total fluxes of each tumor were normalized to time 0 flux, which is defined as the first day of detection after intracranial implant.

### Stable isotope infusions in GBM-bearing mice

At approximately 1-2 weeks before expected death related to brain tumors (∼3 weeks post-implant), intracranial GBM-bearing mice underwent dual catheterizations, with one catheter surgically placed into the jugular vein (for [U^13^C]-glucose administration) and a second catheter placed into the carotid artery (for plasma collection during infusion). Mice were then allowed to recover from surgery for 4-5 days. In studies assessing the influence of RT on metabolism, cannulated mice were anesthetized by 2% isoflurane inhalation and then treated with cranially directed RT at a dose of 8 Gy or sham RT with a lead shield keeping the cranium exposed. Immediately after RT (<5 min), awake and active mice were then administered a bolus dose of [U^13^C]-glucose (0.4 mg/g) followed by a continuous [U^13^C]-glucose infusion (12 ng/g/min) via the intravenous line for a total of 4 h. During infusions, blood was collected periodically via the carotid line into EDTA-coated vials and used to prepare plasma. At the end of infusions, ketamine (50 mg/kg) was administered into the intravenous line rapidly induce anesthesia. Mice were then decapitated, and tissues were extracted on dry ice. To separate orthotopic GBM from normal mouse cortex, we performed microdissection aided by a fluorescent bulb that allowed us to distinguish GFP^+^ tumor from GFP^-^ cortex. All tissues were then immediately (<3 min post-anesthesia) flash-frozen in liquid nitrogen for further analysis.

### Liquid chromatography-mass spectrometry

Flash-frozen tissue samples were homogenized in cold (-80 °C) 80% methanol. For plasma analysis, 100% methanol at -80 °C was added to plasma samples to yield a final methanol concentration of 80%. Insoluble material was then precipitated from all samples by centrifugation at 4 °C, and supernatants containing soluble metabolites were dried by nitrogen purging. Dried metabolites were then reconstituted in 50% methanol for LC-MS analysis. Isotope labeling was determined using an Agilent system consisting of an Infinity Lab II UPLC coupled with a 6545 QTOF mass spectrometer (Agilent Technologies, Santa Clara, CA), and data were analyzed with values corrected for natural isotope abundance using MassHunter Profinder 10.0. We used control samples without ^13^C labeling to ensure that labeled isotopologs from [U^13^C]-glucose-infused mice and patients were not from contaminating species. To determine relative metabolite abundances in tissues and plasma from control or Ser/Gly (-) diet fed mice, samples were prepared as described above and then analyzed with an Agilent 1290 Infinity II LC–6470 Triple Quadrupole tandem mass spectrometer system (Agilent Technologies, Santa Clara, CA). For compound optimization, calibration, and data acquisition, Agilent MassHunter Quantitative Analysis Software version B.08.02 was used.

### MALDI mass spectrometry

Standard microscope slides with mounted tissue were vacuum desiccated for 20 minutes prior to matrix coating. After desiccation, slides were sprayed with NEDC matrix (10 mg/mL, 1:1 ACN:H_2_O) using an M3+ sprayer (HTX Technologies LLC, Chapel Hill, North Carolina, flow rate: 75 µL/min, temperature: 70°C, velocity: 1000 mm/min, track spacing: 1 mm, pattern: crisscross, drying time: 0 sec). Slides were mounted into a MTP Slide Adapter II (Bruker Daltonics, Billerica, MA) before analysis.

MALDI imaging data were acquired using a timsTOF fleX MALDI-2 mass spectrometer (Bruker Daltonics) operating in transmission mode with a 20 µm raster size, acquiring m/z 100–600. The laser (Bruker Daltonics; SmartBeam 3D, 355 nm, 5000 Hz repetition rate) utilized a 16 µm beam scan, resulting in a 20 µm x20 µm ablation area. Taurine was used as a lock mass ([M-H]1-, m/z 124.0074).

Mass spectrometry imaging data were visualized using SCiLS Lab 2023b, with single fractional enrichment, normalized mean enrichment, and fractionalized enrichment images generated in SCiLS Lab using an in-house script utilizing the SCiLS REST API (Bruker Daltonics; version 6.2.114), written in R (version 4.2.2), using RStudio (2022.12.0 Build 353). A segmentation algorithm built into SCiLS Lab was used to create four data-driven regions corresponding to the healthy and GBM tissue in the ^13^C dosed and control tissues (normalization: total ion count, denoising: weak, method: bisecting k-means, metric: Manhattan). Relative isotopologue intensity of these regions was also calculated with another in-house script implemented through the SCiLS REST API.

Tentative annotations were performed using MetaboScape 2023 (Bruker Daltonics) using target lists of known biological molecules generated with the TASQ software (Bruker Daltonics; amino acids, glycolysis, citrate cycle, urea cycle, bile acid, gangliosides) as well as LipidBlast and LIPIDMAPS. The molecular formula of target molecules was used to calculate an accurate mass for each target. Annotations required a mass error of less than 3.0 ppm. In total, 72 features were annotated using this limited list, with annotated peaks having a mass accuracy of 1.1 ppm.

### Metabolic flux analysis

*In vivo* Metabolic Flux Analysis (iMFA) method was developed to estimate the purine and pyrimidine fluxes from isotopologue time-course data. iMFA is described in detail in the supplementary methods. A steady-state IMM-MFA method was used to estimate the serine contribution in human brain tumors, which is also described in detail in the supplementary methods.

### Score of de-novo serine synthesis relative to cortex

A score was defined to compare the relative serine synthesis and uptake between the cortical and tumor tissues. The ratio of de-novo serine synthesis to serine uptake was estimate for each tissue along with the 95% confidence interval of the ratio (Fig S13B). The score was subsequently estimated by dividing the ratio of tumor tissues to that of matched cortical tissue.

### Dietary restriction of serine and glycine

Three days prior to intracranial tumor implantation, mice were placed on either a control diet containing 1.00% serine and 0.99% glycine (TestDiet® Baker Amino Acid Diet, 5CC7), or a modified diet (TestDiet® Modified Baker Amino Acid Diet, 5BJX) containing 0% serine and 0% glycine with all other amino acids adjusted to account for serine and glycine reduction. Mice were then implanted with intracranial tumors and maintained on respective diets for the remainder of experimentation. Tumor growth was monitored by BLI. Once control mice neared humane endpoints, mice were deeply anesthetized, and blood was collected by cardiac puncture into EDTA-coated vials for plasma preparation. Mice were then decapitated, and brains were bisected through tumor tissue. One half of brain was fixed and embedded for histopathologic analysis, which was performed as previously described^9^. The other half of brain underwent rapid GFP^+^-based separation of tumor and cortex as described above. Tissues were then rapidly harvested on dry ice and flash frozen in liquid nitrogen for metabolomic analysis by LC-MS on an Agilent 6470 mass spectrometer as described above.

### Statistical analysis of metabolite enrichment

Statistical analysis was performed either in R or in GraphPad Prism. Data were tested for normal distribution using the D’Agostino normality test (n >= 8) or the Shapiro-Wilk n ormality test (n < 7). For normally distributed data, a t-test was used for two groups, and a one-way ANOVA followed by Tukey HSD was used for more than two groups. For non-normal data, a Mann-Whitney U test was used for two groups. For multiple groups, the Kruskal-Wallis test was used, followed by Mann-Whitney U test with Bonferroni correction for pairwise comparison. To reduce chances of false positives in the differential enrichment analysis, FDR correction was used.

## Data Availability

All data produced in the present study are available upon reasonable request to the authors

## Acknowledgements

We thank Dr. Jann Sarkaria for providing PDX models used in animal studies, the neurosurgical team at the University of Michigan for assisting with human stable isotope infusions, and the patients enrolled in our clinical tracing study. We are grateful to Dr. Ralph DeBerardinis for sharing his advice and expertise in human stable isotope tracing. AJS was supported by the NCI (F32CA260735). DRW was supported by the NCI (K08CA234416; R37CA258346), the NINDS (R01NS129123), the Damon Runyon Cancer Foundation, the Sontag Foundation, the Ivy Glioblastoma Foundation, the Forbes Institute for Cancer Discovery, Alex’s Lemonade Stand Foundation, and the Chad Tough Defeat DIPG foundation. DRW and TSL were supported by NCI P50CA269022. DN was supported by the NCI (R01CA271369). ZW, JF, and NQ were supported by the NIDDK MMPC-*Live* (1U2CDK135066). W.Z was supported by the University of Michigan Medical School Pandemic Research Recovery grant (U083054). Some illustrations were created using BioRender software.

