## Supplement for "Rewiring of cortical glucose metabolism fuels human brain cancer growth"

**MALDI FT-ICR mass spectrometry imaging of clinical specimens**

Clinical specimens, comprising cortex, enhancing, and non-enhancing regions, were sectioned at 10 µm thickness and then thaw-mounted onto indium-tin-oxide (ITO) slides. Serial sections were obtained for hematoxylin and eosin (H&E) staining. A high-resolution image of the entire H&E tissue samples was captured using image stitching technology (Zeiss Observer Z.1, Oberkochen, Germany) with a plan-apochromat lens (20×) equipped with an AxioCam MR3 camera. A matrix solution of 1,5-diaminonaphthalene hydrochloride was prepared at a concentration of 4.3 mg/mL in a mixture of 4.5 parts HPLC-grade water, 5 parts ethanol, and 0.5 parts 1 M HCl (v/v/v). A TM-sprayer (HTX imaging, Carrboro, NC) was used to apply a uniform layer of MALDI matrix, with the following four-pass spray method: a flow rate of 0.09 mL/min, spray nozzle velocity of 1200 mm/min, spray nozzle temperature of 75°C, nitrogen gas pressure at 10 psi, a track spacing of 2 mm, and Mass spectrometry imaging experiments were conducted in negative ion mode using a 15 Tesla SolariX XR FT-ICR MS (Bruker Daltonics, Billerica, MA). The instrument was calibrated with a tune mix solution (Agilent Technologies, Santa Clara, CA) via the electrospray source. Instrumental parameters included a pixel step size of 50 µm, covering the m/z range of 46–300. Each pixel was acquired with 200 shots at a laser power of 19% (arbitrary scale) and a frequency of 1000 Hz. Continuous accumulation of selected ions (CASI) mode was employed, with Q1 set to m/z 180 and an isolation window of 200.

The obtained datasets were preprocessed, visualized, and exported to imZML using SCiLS Lab software (Bruker Daltonics, Billerica, MA). An in-house R pipeline was used to perform per-pixel natural abundance correction and produce fractional enrichment images. The pipeline uses rMSIproc [https://doi.org/10.1093/bioinformatics/btaa142] to import data and enviPat [https://doi.org/10.1021/acs.analchem.5b00941] to compute theoretical MS spectrum for each ^13^C isotopomer. Natural abundance correction is performed by solving a system of linear equations, taking into account the theoretical isotopomer MS ratios and the experimental MS spectrum at each pixel [https://doi.org/10.1002/(SICI)1096-9888(199603)31:3%3C255::AID-JMS290%3E3.0.CO;2-3].

**iMFA at Metabolic Steady State**

Several adaptations were made to a previously published *in vitro* INST-MFA method to make it applicable to *in vivo* data [1]. The model parameters comprised of reaction fluxes (expressed as vector ***v***), the pool sizes of mass-balanced metabolites (expressed as vector ***c***), the isotopologues of input metabolites (**R**), and the fraction of contribution of reactant isotoplogues to the product isotopologues (***f***) used only in pyrimidine model. The vector of model parameters, ***x*** is described in equation 1.

$$\boldsymbol{x}=[\boldsymbol{v},\boldsymbol{c},\boldsymbol{f},\boldsymbol{R}]$$

**Equation 1**

Metabolites inside the model boundary were mass-balanced and are called balanced metabolites. The metabolites outside the model boundary were not mass-balanced and are called input metabolites. Due to the complicated time-dependent nature of *in vivo* metabolite enrichments, this demarcation between input metabolites and balanced metabolites was important to establish the model. A set of linear mass balance equations were used to describe overall mass balance. The sum of the isotopologues of the input metabolites was constrained to 1. Equation 2 describes the linear constraint equations. ***S*** is the stoichiometric matrix for a model with *m* balanced metabolites and *n* reactions. ***L*** is the linear constraints on the sum of input metabolite isotopologues, corresponding to *p* labeled input metabolites and *r* total isotopologues.

$$\left[ {S_{m\times n} \atop0_{p\times n}} {0_{m\times m} \atop0_{p\times m}} {0_{m\times q} \atop0_{p\times q}} {0_{m\times r} \atop L_{p\times r}} \right] x= \left[ {0_{m\times1} \atop1_{p\times1}} \right]$$

**Equation 2**

The time-dependent fractional isotopologue enrichment (MIDs) of balanced metabolites is described by a set of ordinary differential equations (ODEs, equation 3). The rate of change of the isotopologue *d* of metabolite *i* (*M_i,d_)* is described by applying mass balance on the isotopologue.

$$\frac{dM_{i,d}}{dt}= \frac{1}{c_{i}}.\left( \sum_{\begin{aligned} j=1 \\ S_{ij}>0 \end{aligned}}^{n} S_{ij}.v_{j}\left( \prod_{\begin{aligned} k \\ \sum q=d \\ S_{kj}<0 \end{aligned}} M_{k,q} \right) + \sum_{\begin{aligned} j=1 \\ S_{ij}<0 \end{aligned}}^{n} S_{ij}.v_{j}.M_{i,d} \right)$$

**Equation 3**

An objective function was minimized to solve the model and estimate the optimal parameters (equation 4) [2]. The objective function (*obj*) was calculated as the sum of square of the differences between the measured values of isotope enrichments (*M_expt_*) and the isotopic enrichments simulated by the model (*M_sim_*) divided by the standard deviation of the experimental measurements (*SD_expt_*). When the metabolite pool sizes were known, the difference between the known and simulated pool sizes were included in the objective function (*c_expt_ - c_sim_*).

$$obj= \sum\left( \frac{M_{expt}-M_{sim}}{{SD}_{expt}} \right)^{2}+ \sum\left( \frac{c_{expt}-c_{sim}}{{SD}_{expt}} \right)^{2}$$

**Equation 4**

To estimate the fluxes and pool sizes, an initial parameter vector was selected randomly and the ODEswere solved to estimate the objective function. The objective function was minimized subject to linear constraints and parameter bounds. The optimization was performed in MATLAB with the Artlelys Knitro toolbox [3]. The solver *ode15s* was used to solve ODEs as Initial Value Problems (IVPs). All metabolites were unlabeled at time *t=0*. The optimization was performed for 100 randomly generated initial parameter vectors and the chi-square-goodness-of-fit test was used to select the optimal parameter vector at 95% confidence [4]. When the objective function was higher than the chi-square threshold, the parameter space with the lowest objective value was selected. A previously described approach was used to determine the 95% confidence intervals for the estimated fluxes [4].

**Dynamic iMFA**

Previously described DMFA approaches were adapted to apply them to *in vivo* data [5,6]. The data for radiation-induced time-dependent change in metabolite pool sizes were incorporated into the model along with the MID-time profiles. The time-course flux profiles were parametrized by expressing them as B-splines (equation 5). B-spline is a parametric function that can be used to fit data without assuming a functional relationship between the input and output variables. It comprises of multiple polynomial segments joined together via ‘control points’. These control points control the shape of the b-spline curve and are hyperparameters in the model. Another hyperparameter is the b-spline order, which is equal to *d+1*, *d* being the degree of polynomials used to construct the b-spline [5].

$$\boldsymbol{v}(t)=\boldsymbol{CP}*\boldsymbol{N}(t)$$

**Equation 5**

The parameter vector comprised of the metabolite pool size at time *t=0* (***c_0_***), and the b-spline parameters expressed as vector ***cp*** and the isotopologues of input metabolites (**R**) (equation 6). The rate of change of metabolite pool size was expressed as a function of the stoichiometric matrix *S* and the time-dependent flux vector ***v****(t)* (equation 7).

$$\boldsymbol{x}=[\boldsymbol{cp},\boldsymbol{c}_{\boldsymbol{0}},\boldsymbol{R}]$$

**Equation 6**

$$\frac{d\boldsymbol{c}}{dt}=\boldsymbol{S}*\boldsymbol{v}(t)$$

**Equation 7**

The objective function from the steady state iMFA model was modified to include the terms for time-dependent metabolite pool size (equation 8). Only the relative time-dependent pool size could be measured experimentally. Hence, the relative change of the metabolite pool size at time *t* (*c_t_*) relative to the pool size at time zero (*c_0_*) was used in the objective function. The initial metabolite pool size values were the same as those used in the steady state model. The relative time-dependent concentration profiles are provided in figure S10B.

$$obj= \sum\left( \frac{M_{expt}-M_{sim}}{{SD}_{expt}} \right)^{2}+ \sum\left( \frac{c_{0,expt}-c_{0,sim}}{{SD}_{expt}} \right)^{2} + \sum\left( \frac{\left( \frac{c_{t}}{c_{0}} \right)_{expt}- \left( \frac{c_{t}}{c_{0}} \right)_{sim}}{{SD}_{expt}} \right)^{2}$$

**Equation 8**

To estimate the flux profiles, an initial parameter vector was randomly selected, and the ODEs were solved to minimize the objective function as done for the steady state model. The flux values at *t=0* were constrained to the 95% confidence intervals estimated from the steady state model. All metabolites were unlabeled at time *t=0*. The optimization was performed for 100 randomly generated initial parameter vectors and the parameter vector with the lowest objective value was selected. To estimate the 95% confidence intervals, the parameter bounds were estimated as done for the steady state model. The determined parameter bounds were used to create the time-course flux profiles corresponding to the 95% confidence interval [7]. All the acceptable flux profiles were recorded and the minimum and maximum flux values at a certain time were reported as the 95% flux bound.

The b-spline hyperparameter selection was performed prior to parameter optimization. B-splines of orders 2,3, and 4 were tested and a spline of order 3 (quadratic b-spline) was selected after qualitative analysis of the results. A randomized approach was used to select the b-spline control points. All flux profiles were assumed to have the same control points. Control points in the range of [0.2,0.8] were tested at intervals of 0.05. Each time, one position was selected, and parameter optimization was performed for the same initial parameter vector. The control point with the lowest objective was recorded. To optimize the placement of a second control point, the same strategy was used. The second control point was placed at a minimum distance of 0.2 from the first control point. The procedure was repeated for 100 initial guesses. For both the GBM and normal cortex samples, the addition of a second control point did not reduce the minimum objective value for the 100 optimizations. Hence, one control point was used to simulate both conditions. Based on the model fit, 0.3 was selected as the control point for GBM because of a lower mean objective value and central position. 0.65 was selected for the normal cortex.

**Metabolic Model of the Purine Pathway**

The metabolic model of the purine synthesis pathway was curated on the basis of experimental data and current literature. The KEGG database was referenced for a list of reactions and the associated enzymes [8]. Data from the human proteome atlas was used to remove the reactions whose enzymes have low expression in GBM [9]. All the reactions were assumed to be unidirectional and further simplifications were made based on the available experimental data. The net fluxes were assumed to be in the direction of purine synthesis. Further, IMP was assumed to directly produce GMP and the intermediate XMP was excluded from the model because the enrichment data for XMP was not available. All the ribose units (R5P, R1P, PRPP) were assumed to have the same enrichment. The final model comprised of 15 reactions and 23 metabolites (figure S13A, tables S1, S2). Six purine metabolites were inside the model boundary and were mass balanced. The rest of the metabolites are substrates in the purine pathway and outside the model boundary. The metabolites adenosine and hypoxanthine were assumed to be unlabeled based on experimental data. The enrichment data for guanine was not available and it was assumed to be produced mainly from the degradation of old DNA and RNA units and hence was unlabeled in the model. The cellular carbon dioxide pool was also assumed to be unlabeled. The enrichment of methyl units cannot be measured experimentally, and the values were estimated from the enrichments of serine and glycine.

**Table S1:** List of model reactions and associated flux bounds.

Reaction type irreversible refers to reactions that proceed only in one direction. Reaction type sink refer to the reactions that consume the metabolite and are not included in the model. Flux bounds were set according to the experimental data. AMP: adenosine monophosphate; C-THF: 5-methytetrahydrofolate; CO_2_: carbon dioxide; GLY: glycine; IMP: inosine monophosphate; GDP: guanosine diphosphate; GMP: guanosine monophosphate; R5P: ribose-5-phosphate

| **Reaction No.** | **Reaction** | **Reaction Type** | **Flux Lower Bound (pmol/mg.hr)** | **Flux Upper Bound**  **(pmol/mg.hr)** |
| --- | --- | --- | --- | --- |
| 1 | R5P + GLY + 2C-THF + CO_2_ == IMP | Irreversible | 0.01 | 300 |
| 3 | IMP == INOSINE | Irreversible | 0.01 | 300 |
| 2 | R5P + HYPOXANTHINE == IMP | Irreversible | 0.01 | 300 |
| 4 | R5P + HYPOXANTHINE == INOSINE | Irreversible | 0.01 | 300 |
| 5 | INOSINE == 0 | Sink | 0.01 | 300 |
| 6 | IMP == GMP | Irreversible | 0.01 | 300 |
| 7 | R5P + GUANINE == GMP | Irreversible | 0.01 | 300 |
| 8 | GMP == GUANOSINE | Irreversible | 0.01 | 300 |
| 9 | R5P + GUANINE == GUANOSINE | Irreversible | 0.01 | 300 |
| 10 | GUANOSINE == 0 | Sink | 0.01 | 300 |
| 11 | GMP == GDP | Irreversible | 0.01 | 300 |
| 12 | GDP == 0 | Sink | 0.01 | 300 |
| 13 | IMP == AMP | Irreversible | 0.01 | 300 |
| 14 | ADENOSINE == AMP | Irreversible | 0.01 | 300 |
| 15 | AMP == 0 | Sink | 0.01 | 300 |

**Table S2:** List of metabolites included in the model.

Input metabolites are not mass balanced. Some input metabolites had zero to low isotopic enrichment and were considered unlabeled.

| **Metabolite No.** | **Metabolite** | **Metabolite Type** | **No. of Carbon Atoms** |
| --- | --- | --- | --- |
| 1 | Ribose-5-Phosphate (R5P) | Input | 5 |
| 2 | Glycine (GLY) | Input | 2 |
| 3 | Carbon Dioxide (CO_2_) | Input (unlabeled) | 1 |
| 4 | 5-Methyltetrahydrofolate (C-THF) | Input | 1 |
| 5 | Inosine Monophosphate (IMP) | Mass-Balanced | 10 |
| 6 | Inosine | Balanced | 10 |
| 7 | Hypoxanthine | Input (unlabeled) | 5 |
| 8 | Guanine | Input (unlabeled) | 5 |
| 9 | Guanosine Monophosphate (GMP) | Mass-Balanced | 10 |
| 10 | Guanosine Diphosphate (GDP) | Mass-Balanced | 10 |
| 11 | Guanosine | Mass-Balanced | 10 |
| 12 | Adenosine Monophosphate (AMP) | Mass-Balanced | 10 |
| 13 | Adenosine | Input (unlabeled) | 10 |

To estimate the methyl unit enrichment, a pseudo-steady state assumption was applied. Serine, glycine, and the methyl enrichments were assumed to be in equilibrium at any given time. The corresponding equations for the metabolite isotopomers are represented in table S3 (SER: serine; GLY: glycine; ME: Me-THF; the number in subscript corresponds to the presence (1) or absence (0) of ^13^C carbon at the position). The variance-weighted sum-of-squared residuals was calculated between the experimental MID values of serine and glycine and the model estimated values. The optimization was performed from 10 initial guesses. To estimate the error in the estimated methyl enrichment, gaussian noise was added to the experimental values and the optimization was repeated 200 times.

**Table S3:** Equations to estimate methyl unit enrichment

| S. No. | Equation |
| --- | --- |
| 1 | ${SER}_{000}={GLY}_{00}{ME}_{0}$ |
| 2 | ${SER}_{001}={GLY}_{00}{ME}_{1}$ |
| 3 | ${SER}_{100+010}={GLY}_{10+01}{ME}_{0}$ |
| 4 | ${SER}_{101+011}={GLY}_{10+01}{ME}_{1}$ |
| 5 | ${SER}_{110}={GLY}_{11}{ME}_{0}$ |
| 6 | ${SER}_{111}={GLY}_{11}{ME}_{1}$ |
| 7 | ${ME}_{0}+{ME}_{1}=1$ |

**Metabolic Model of the Pyrimidine Pathway**

Pyrimidine model was implemented based on our available enrichment data for pyrimidines, the active reactions in KEGG database, and the enzymes that have high expression in GBM by checking the human protein atlas [8,9]. The pyrimidine model consists of three reactions assumed to be unidirectional toward synthesis of UMP and listed in Table S4 and depicted in figure S13B. Flux bounds were selected based on our observation of optimization space and were relaxed to have no overlap with the flux confidence intervals.

**Table S4**: List of pyrimidine model reactions and associated flux bounds.

Reaction type irreversible refers to reactions that proceed only in one direction. Reaction type sink refer to the reactions that consume the metabolite and are not included in the model. Flux bounds were set according to the experimental data. UMP: uridine monophosphate; CO_2_: carbon dioxide; R5P: ribose-5-phosphate; ASP: aspartate.

| **Reaction No.** | **Reaction** | **Reaction Type** | **Flux Lower Bound (pmol/mg.hr)** | **Flux Upper Bound**  **(pmol/mg.hr)** |
| --- | --- | --- | --- | --- |
| 1 | R5P + ASP + CO_2_ == UMP + CO2 | Irreversible | 0.01 | 100 |
| 2 | URIDINE == UMP | Irreversible | 0.01 | 200 |
| 3 | UMP == 0 | Sink | 0.01 | 300 |

**Table S5:** List of metabolites included in the pyrimidine model.

Input metabolites are not mass balanced. Some input metabolites had zero to low isotopic enrichment and were considered unlabeled.

| **Metabolite No.** | **Metabolite** | **Metabolite Type** | **No. of Carbon Atoms** |
| --- | --- | --- | --- |
| 1 | Ribose-5-Phosphate (R5P) | Input | 5 |
| 2 | Aspartate (ASP) | Input | 4 |
| 3 | Carbon Dioxide (CO_2_) | Input (unlabeled) | 1 |
| 4 | Uridine | Input | 9 |
| 5 | Uridine Monophosphate (UMP) | Mass-Balanced | 9 |

The input metabolites of our model include R5P, aspartate, CO_2_, and uridine (Table S5). We assumed that all ribose units including R5P, PRPP, and R1P share the same enrichment profiles. The only metabolite that is inside the model is UMP and we wrote the mass isotopologue balance on it (equation 9).

$$\frac{dM_{UMP, i}}{dt}=\frac{1}{c_{UMP}}\left( v_{salvage}.M_{URIDINE,i}+v_{de novo}.M_{R5P,j}.\tilde{M}_{ASP, k}-v_{UMP_{tot}}.M_{UMP,i} \right);j+k=i$$

**Equation 9**

On the left side of the equation 9, the rate of change of UMP isotopologues is calculated. $c_{UMP}$ denotes the concentration of UMP. The first term shows that UMP gets labeling from the salvage flux ($v_{salvage}$) through uridine MIDs ($M_{URIDINE,i}$ where $i$ denotes the M+i enrichment). The second term show UMP gets labeling from the de novo UMP flux ($v_{de novo}$) through R5P ($M_{R5P,j}$) and aspartate ($\tilde{M}_{ASP, k}$) MIDs. In this model, we assumed that CO_2_ is unlabeled. The produced UMP in de novo synthesis consists of nine carbons in which five carbons come from R5P to structure the ribose unit of UMP. The remaining four carbons in the uracil ring come from unlabeled CO_2_ pool (2^nd^ position), and aspartate (4^th^, 5^th^, and 6^th^ positions). UMP de novo synthesis is a decarboxylation reaction and the carbon leaving the system as CO_2_ might be labeled if the first carbon of aspartate is labeled. Therefore, we defined a new isotopologue distribution for aspartate ($\tilde{M}_{ASP, k}$) that only contributes to the UMP labeling. Table S6 shows aspartate isotopomers that correspond to each isotopologue of aspartate ($M_{ASP, M+i}$) that can enrich UMP in the same way. To do so, three isotopomer fractions were defined ($f_{1}, f_{2}, f_{3})$. For instance, suppose one labeled carbon in UMP comes from aspartate (${\tilde{\boldsymbol{M}}}_{\boldsymbol{ASP,M+1}}$), since the labeling of the first carbon of aspartate does not affect the labeling of UMP, the following isotopomers of aspartate can produce one labeled carbon in UMP: 0001, 0010, 0100, 1001, 1010, and 1100. The defined fractions $f_{i}$ corresponded to M+i isotopologue helped us to separate the isotopomers to isotopomers with labeled produced CO_2_ and with unlabeled produced CO_2_. For example, $f_{1}$ is the ratio of isotopomer 1000 to all isotopomers of M+1 (0001, 0010, 0100, 1000). To constrain the lower and upper bounds of fractions, we used the isotopomer distributions in mouse and human tumors reported in literature [10]. Hence, we set the constraints to [0.1, 0.4], [0.2, 0.6], and [0, 1] for $f_{1}, f_{2},$ and $f_{3}$, respectively. To combine the probability of production of UMP M+i isotopologues from aspartate and R5P, we multiplied M+j of R5P ($M_{R5P,j}$) to M+k of newly defined aspartate isotopologues ($\tilde{M}_{ASP, k}$) where j + k = i. The third term in equation 9 represents the consumption of UMP in other reactions outside of the boundaries of the model or the total UMP synthesis.

**Table S6:** Contribution of aspartate labeling in UMP labeling.

| **Newly defined aspartate isotopologues contributed to UMP labeling** | **Aspartate isotopomers that produce unlabeled CO_2_** | **Aspartate isotopomers that produce labeled CO_2_** |
| --- | --- | --- |
| ${\tilde{\boldsymbol{M}}}_{\boldsymbol{ASP,M+0}}\boldsymbol{=}\boldsymbol{M}_{\boldsymbol{ASP, M+0}}\boldsymbol{+}\boldsymbol{f}_{\boldsymbol{1}} \boldsymbol{M}_{\boldsymbol{ASP,M+1}}$ | $0000$ | $f_{1}:1000$ |
| ${\tilde{\boldsymbol{M}}}_{\boldsymbol{ASP,M+1}}\boldsymbol{=}\left( \boldsymbol{1-}\boldsymbol{f}_{\boldsymbol{1}} \right) \boldsymbol{M}_{\boldsymbol{ASP, M+1}}\boldsymbol{+}\boldsymbol{f}_{\boldsymbol{2}} \boldsymbol{M}_{\boldsymbol{ASP,M+2}}$ | $\left( 1-f_{1} \right):0001, 0010, 0100$ | $f_{2}:1001, 1010, 1100$ |
| ${\tilde{\boldsymbol{M}}}_{\boldsymbol{ASP,M+2}}\boldsymbol{=}\left( \boldsymbol{1-}\boldsymbol{f}_{\boldsymbol{2}} \right) \boldsymbol{M}_{\boldsymbol{ASP, M+2}}\boldsymbol{+}\boldsymbol{f}_{\boldsymbol{3}} \boldsymbol{M}_{\boldsymbol{ASP,M+3}}$ | $\left( 1-f_{2} \right):0011, 0101, 0110$ | $f_{3}:1011, 1101, 1110$ |
| ${\tilde{\boldsymbol{M}}}_{\boldsymbol{ASP,M+3}}\boldsymbol{=}\left( \boldsymbol{1-}\boldsymbol{f}_{\boldsymbol{3}} \right) \boldsymbol{M}_{\boldsymbol{ASP, M+3}}\boldsymbol{+}\boldsymbol{M}_{\boldsymbol{ASP,M+4}}$ | $\left( 1-f_{3} \right):0111$ | 1111 |

**Metabolite Pool Sizes**

Purine and pyrimidine pool sizes for mouse brain were obtained from literature, when available. Only the values reported with the measurement standard deviation were included in the objective function. When the measurement error was not available, the reported value was only used to set the bounds for the corresponding pool size parameter. Experimental data was used to estimate the pool sizes for GBM tissue. The average relative metabolite ion count between GBM and brain tissue was calculated and multiplied by the pool size in the brain. Standard error propagation techniques were used to estimate the standard deviation of the GBM pool sizes. GMP was not detected in the brain tissue but was detected in the GBM tissue. Hence, the bounds for GMP concentration were set to be higher than the pool size in the brain. The pool size data and source literature are reported in table S7.

**Table S7:** Purine and pyrimidine pool sizes used in metabolic model.

| **Metabolite** | **Pool Size in Mouse Brain** (Mean ± SD, pmol/mg tissue) | **Pool Size in GBM**  (Mean ± SD, pmol/mg tissue) | **Additional Bounds on Pool Size**  (pmol/mg tissue) |
| --- | --- | --- | --- |
| GDP | 182 .1± 8.2^[11]^ | 59.0 ± 22.1 |  |
| Inosine | 126.7 ± 18.3^[12]^ | 94.1 ± 26.8 |  |
| AMP | 172 ± 6.0^[11]^ | 95.4 ± 48.9 |  |
| Guanosine | 245.8 ± 31.6^[12]^ | 70.5 ± 22.0 |  |
| GMP | 157 ± 40^[13]^ |  | GBM: [172,304] |
| IMP | ~200^[14]^ |  | Brain: [125,300]  GBM: [30,70] |
| UMP | 3.16 ± 1.26^[15]^ | 2.31 ± 1.10 |  |

**Enrichment of Input Metabolites**

The time-course isotopologue abundances of input metabolites are required to apply the metabolic model. The time-course enrichment of metabolites is complex in *in vivo* models and cannot be described by exact mathematical functions. A linear piecewise function was fit to the experimental time-point data to estimate the enrichment-time relationship of input metabolites. Equation 10 describes the calculation of the slope of the isotopologue *R_i_* between time points *t_m+1_* and *t_m_*. A linear function was selected because it requires the least assumptions and does not overfit the data. To account for the uncertainty in the experimental measurements, the input isotopologue abundances were allowed to vary within one standard deviation of the experimentally measured mean value.

$$\frac{dR_{i}(t)}{dt}= \left\{ \begin{aligned} \frac{R_{i}\left( t_{2} \right)-R\left( t_{1} \right)}{t_{2}- t_{1}} t_{1}\leq t<t_{2} \\ \ldots\\ \ldots\\ \frac{R_{i}\left( t_{m+1} \right)-R_{i}\left( t_{m} \right)}{t_{m+1}- t_{m}}t_{m}\leq t<t_{m+1} \end{aligned} \right.$$

**Equation 10**

**MFA of Serine Contribution in Human Tumors**

The model comprised of three compartments which correspond to the cortex, enhancing tumor, and non-enhancing tumor (figure 6H). Each compartment had its own 3PG pool which was used for *de-novo* serine synthesis within the compartment. The compartments were linked by a common input of external serine from circulation. A source of unlabeled serine was included in the model since serine might be derived from protein breakdown. Autophagy has been shown to be a source of serine in *in vitro* models of glioma [16]. Further, *de-novo* serine synthesis is rate limited by the levels of the PHGDH enzyme, and serine may not be in isotopic equilibrium with 3PG. An unlabeled serine source helped correct for this effect. The net serine input to the model from *de-novo* synthesis, external serine uptake, and the unlabeled serine from recycle was assumed to be 1 unit. Forward and reverse SHMT fluxes and the fluxes of glycine to 5,10-methylene-THF were included in each compartment. Serine was assumed to be the only source of glycine and one-carbon unit. This is in accordance with previous findings which show that glycine is mainly derived from serine in the brain [17,18]. Further, serine has been described as a major source of glycine and one-carbon units in gliomas [16,19]. The enrichment of externally available serine was also assumed to be the same as the serine in circulation. Further, we assumed that tissues are homogenous and did not consider any cell-cell metabolic interactions in the serine-glycine pathway. These assumptions were important to reduce the model complexity and prevent the model from becoming highly underdetermined. Because of the various unknown factors, we only analyzed and reported the relative contributions of serine synthesis and uptake pathways that would yield labeled serine and did not analyze the absolute values. Sink fluxes were also included for serine, glycine, and formyl-THF to account for consumption not included in the model. The list of reactions and the associated carbon transitions are provided in table S8.

The model parameters *x* comprised of the fluxes *v* and the known IDV values *D* of 2PG/3PG, serine, and glycine in the three compartments. The MID of plasma serine was also included in the model parameters (equation 11). The fluxes in each compartment were mass balanced, which was described through the stoichiometric matrix ***S****_m×n_* corresponding to *m* metabolites and *n* reactions (equation 12). The external serine was not included in the mass balance. To make sure that the sum of all IDVs of a certain metabolite is always equal to 1, the sums of the IDVs of the external serine and the three tissue 3PG were constrained to 1. This added 4 equations to the model (one for external serine, 3 for 3PG in the three tissue compartments). These equations were represented by the matrix ***L****_4×d_* corresponding to *d* total IDV values in the model. The three equations constraining the new serine input to 1 were also included in the linear balance equations and are represented by the matrix ***N****_3×n_*.

$$\boldsymbol{x}=[\boldsymbol{v},\boldsymbol{D}]$$

**Equation 11**

$$\left[ \begin{matrix} \boldsymbol{S}_{m\times n} & \boldsymbol{0}_{m\times d} \\ \boldsymbol{N}_{3\times n} & \boldsymbol{0}_{3\times d} \\ \boldsymbol{0}_{4\times n} & \boldsymbol{L}_{4\times d} \end{matrix} \right]\boldsymbol{x}=\left[ \begin{matrix} \boldsymbol{0}_{n\times1} \\ \boldsymbol{1}_{7\times1} \end{matrix} \right]$$

**Equation 12**

The IMM method was used to formulate the isotopic mass balance equations (equation 13) [20]. These equations were used to add non-linear constraints to the model. Additional linear constraints were applied to avoid the model converging to a trivial solution. The parameters were optimized to minimize the objective function, which was defined to minimize the differences between the experimentally measured MIDs and the MIDs simulated by the model (equation 14). The difference between the experimental and simulated values was normalized to the experimental standard deviation to account for experimental variation. An L2 regularization term for the flux parameters was also included in the objective function because the model was underdetermined, i.e. we had more unknown parameters than the number of known values. The value of the regularization parameter was set to 0.1. To solve the model, local optimization was performed from 200 randomly selected initial points. Knitro toolbox was used for the optimization in MATLAB. The solution with the lowest objective value was selected. To estimate the 95% confidence intervals, gaussian noise was added to the experimental data and the optimization was repeated 1000 times. The distribution of the resulting solutions was used to determine the confidence intervals.

$$\sum_{\begin{aligned} j=1 \\ S_{ij}>0 \end{aligned}}^{n} S_{ij}.v_{j}\left( \prod_{\begin{aligned} k \\ \sum q=d \\ S_{kj}<0 \end{aligned}} M_{k,q} \right) + \sum_{\begin{aligned} j=1 \\ S_{ij}<0 \end{aligned}}^{n} S_{ij}.v_{j}.M_{i,d}=0$$

**Equation 13**

$$obj= \sum\left( \frac{\sum y_{sim}-M_{expt}}{{SD}_{expt}} \right)^{2}+0.1*{||[v,y]||}_{2}$$

**Equation 14**

**Table S8:** List of reactions in the MFA model.

SER: serine; GLY: glycine; 3PG: 3-phosphoglycerate; Me-THF: 5,10-methylene-THF

| S. No. | Reaction | Carbon Transition |
| --- | --- | --- |
| 1 | SER_plasma_ == SER_cortex_ | abc == abc |
| 2 | 3PG_cortex_ == SER_cortex_ | abc == abc |
| 3 | SER_unlabeled_ == SER_cortex_ | abc == abc |
| 4 | SER_cortex_ == GLY_cortex_ + Me-THF_cortex_ | abc == ab + c |
| 5 | GLY_cortex_ + Me-THF_cortex_ == SER_cortex_ | ab + c == abc |
| 6 | GLY_cortex_ == Me-THF_cortex_ + CO_2_ | ab == b + a |
| 7 | SER_cortex_ == 0 |  |
| 8 | GLY_cortex_ == 0 |  |
| 9 | Me-THF_cortex_ == 0 |  |
| 10 | SER_plasma_ == SER_enhancing_ | abc == abc |
| 11 | 3PG_enhancing_ == SER_enhancing_ | abc == abc |
| 12 | SER_unlabeled_ == SER_enhancing_ | abc == abc |
| 13 | SER_enhancing_ == GLY_enhancing_ + Me-THF_enhancing_ | abc == ab + c |
| 14 | GLY_enhancing_ + Me-THF_enhancing_== SER_enhancing_ | ab + c == abc |
| 15 | GLY_enhancing_ == Me-THF_enhancing_ + CO_2_ | ab == b + a |
| 16 | SER_enhancing_== 0 |  |
| 17 | GLY_enhancing_ == 0 |  |
| 18 | Me-THF_enhancing_ == 0 |  |
| 19 | SER_plasma_ == SER_nonenhancing_ | abc == abc |
| 20 | 3PG_nonenhancing_ == SER_nonenhancing_ | abc == abc |
| 21 | SER_unlabeled_ == SER_nonenhancing_ | abc == abc |
| 22 | SER_nonenhancing_ == GLY_nonenhancing_ + Me-THF_nonenhancing_ | abc == ab + c |
| 23 | GLY_nonenhancing_ + Me-THF_nonenhancing_== SER_nonenhancing_ | ab + c == abc |
| 24 | GLY_nonenhancing_ == Me-THF_nonenhancing_ + CO_2_ | ab == b + a |
| 25 | SER_nonenhancing_== 0 |  |
| 26 | GLY_nonenhancing_ == 0 |  |
| 27 | Me-THF_nonenhancing_ == 0 |  |

**Table S9:** Estimated purine fluxes for GBM tissue. Flux bounds represent 95% confidence intervals.

| **Reaction No.** | **Reaction** | **Flux**  **(pmol/mg.hr)** | **Lower Bound**  **(pmol/mg.hr)** | **Upper Bound**  **(pmol/mg.hr)** |
| --- | --- | --- | --- | --- |
| 1 | R5P + GLY + 2C-THF + CO_2_ == IMP | 8.0 | 6.9 | 8.0 |
| 2 | IMP == INOSINE | 77.2 | 58.8 | 93.7 |
| 3 | R5P + HYPOXANTHINE == IMP | 250.4 | 214.7 | 263.1 |
| 4 | R5P + HYPOXANTHINE == INOSINE | 87.9 | 77.8 | 99.0 |
| 5 | INOSINE == 0 | 165.1 | 102.9 | 190.7 |
| 6 | IMP == GMP | 96.9 | 67.5 | 120.2 |
| 7 | R5P + GUANINE == GMP | 81.1 | 79.5 | 96.6 |
| 8 | GMP == GUANOSINE | 0.2 | 0.2 | 0.2 |
| 9 | R5P + GUANINE == GUANOSINE | 297.2 | 267.5 | 300.0 |
| 10 | GUANOSINE == 0 | 297.4 | 240.9 | 300.0 |
| 11 | GMP == GDP | 177.8 | 125.6 | 277.0 |
| 12 | GDP == 0 | 177.8 | 125.6 | 277.0 |
| 13 | IMP == AMP | 84.3 | 72.1 | 92.7 |
| 14 | ADENOSINE == AMP | 27.3 | 27.3 | 43.7 |
| 15 | AMP == 0 | 111.6 | 74.1 | 183.7 |

**Table S10:** Estimated purine fluxes for normal cortex tissue. Flux bounds represent 95% confidence intervals.

| **Reaction No.** | **Reaction** | **Flux**  **(pmol/mg.hr)** | **Lower Bound**  **(pmol/mg.hr)** | **Upper Bound**  **(pmol/mg.hr)** |
| --- | --- | --- | --- | --- |
| 1 | R5P + GLY + 2C-THF + CO_2_ == IMP | 1.0 | 0.1 | 3.4 |
| 2 | IMP == INOSINE | 1.6 | 0.1 | 61.5 |
| 3 | R5P + HYPOXANTHINE == IMP | 98.3 | 71.7 | 300.0 |
| 4 | R5P + HYPOXANTHINE == INOSINE | 24.3 | 13.5 | 71.0 |
| 5 | INOSINE == 0 | 25.9 | 17.6 | 46.5 |
| 6 | IMP == GMP | 7.9 | 0.1 | 65.3 |
| 7 | R5P + GUANINE == GMP | 93.6 | 75.8 | 300.0 |
| 8 | GMP == GUANOSINE | 17.1 | 1.9 | 41.4 |
| 9 | R5P + GUANINE == GUANOSINE | 45.4 | 20.4 | 81.5 |
| 10 | GUANOSINE == 0 | 62.6 | 37.1 | 109.0 |
| 11 | GMP == GDP | 84.3 | 63.6 | 300.0 |
| 12 | GDP == 0 | 84.3 | 63.6 | 300.0 |
| 13 | IMP == AMP | 89.8 | 71.5 | 285.4 |
| 14 | ADENOSINE == AMP | 0.0 | 0.0 | 16.2 |
| 15 | AMP == 0 | 89.8 | 76.3 | 300.0 |

**Table S11:** Estimated pyrimidine fluxes for GBM tissue. Flux bounds represent 95% confidence intervals.

| **Reaction No.** | **Reaction** | **Flux**  **(pmol/mg.hr)** | **Lower Bound**  **(pmol/mg.hr)** | **Upper Bound**  **(pmol/mg.hr)** |
| --- | --- | --- | --- | --- |
| 1 | R5P + ASP + CO_2_ == UMP + CO2 | 7.2 | 3.1 | 12.6 |
| 2 | URIDINE == UMP | 52.3 | 33.0 | 149.1 |
| 3 | UMP == 0 | 59.6 | 25.6 | 94.8 |

**Table S12:** Estimated pyrimidine fluxes for normal cortex tissue. Flux bounds represent 95% confidence intervals.

| **Reaction No.** | **Reaction** | **Flux**  **(pmol/mg.hr)** | **Lower Bound**  **(pmol/mg.hr)** | **Upper Bound**  **(pmol/mg.hr)** |
| --- | --- | --- | --- | --- |
| 1 | R5P + ASP + CO_2_ == UMP + CO2 | 1.5 | 1.3 | 1.8 |
| 2 | URIDINE == UMP | 54.3 | 53.7 | 78.3 |
| 3 | UMP == 0 | 55.8 | 48.2 | 62.6 |

**Supplementary Figure Legends**

**Figure S1.** Circulating lactate enrichment and specimen histology. (A). Time course of M+3 lactate in plasma from patients infused with [U^13^C]-glucose. (B). Time course of M+3 lactate in plasma from orthotopic GBM bearing mice (n between 2 and 10 for each time point) infused with [U^13^C]-glucose. Data are shown as average ± standard deviation. (C) Representative hematoxylin and eosin stains of tissues resected from our cohort of patients. (D) Percent tumor content in tissues from each patient was defined by a clinical neuropathologist (SV). Data are shown as average ± standard deviation, n=8 patients. *p<0.05. **p<0.01. ns, not significant. Abbreviations: NAA, N-acetylaspartate.

**Figure S2.** Altered metabolite abundances in glioma compared to normal cortex. (A) Levels of NAA in cortical tissue and tumor tissue (enhancing and non-enhancing) from human glioma patients undergoing surgical resection. (B-D) Volcano plots of metabolite abundance determined by LC-MS were used to compare fold change in tumor metabolite levels compared to cortical metabolite levels as follows: (B) enhancing tumor compared to cortex in patients, (C) non-enhancing tumor compared to cortex in patients, and (D) GBM compared to cortex in orthotopic GBM bearing mice. **p<0.01. Abbreviations: NAA, N-acetylaspartate; NAAG, N-acetylaspartylglutamate; GMP, guanosine monophosphate.

**Figure S3.** Isotopic enrichment of UDP-glucose in cortex and glioma. (A) UDP-glucose M+6, predominantly derived from M+6 glucose (left), and UDP-glucose fractional enrichment, accounting for all ^13^C isotopologues (right). N=7 mice. (B) Normalized (to plasma m+6 glucose on a per-patient basis) M+6 UDP-glucose enrichment in tissues from glioma patients infused with [U^13^C]-glucose. Data are shown as average ± standard deviation. *p<0.05. ns, not significant.

**Figure S4.** Spatially defined isotope labeling in mouse brain and GBM. (A) Hematoxylin and eosin staining of brains from orthotopic GBM bearing mice intraperitoneally injected with either vehicle (as a negative control for panels B-G) or [U^13^C]-glucose. MALDI-MS was used to determine ^13^C enrichment of (B) lactate, (C) aspartate, (D) GABA, (E) glutamate, (F) glutamine, and (G) AMP M+5. Tissue maximum is set to 100%. Abbreviations: GABA, gamma-aminobutyric acid; AMP, adenosine monophosphate.

**Figure S5.** Spatially defined isotope labeling in human cortex and glioma. The indicated tissue types resected from brain cancer patients receiving either [U^13^C]-glucose infusion (patients 1-8) or no infusion (unlabeled 1, 2) were assessed by MALDI-MS for ^13^C isotope labeling of (A) malate, (B) glutamate, and (C) glutamine.

**Figure S6.** Enrichment of ^13^C-labeled metabolites quantified by spatial MALDI-MS. ^13^C labeling of (A) malate, (B) glutamate, and (C) glutamine for all data points from spatial MALDI-MS scans of tissues resected from glioma patients (shown in Fig S4). Patients 1-8 were infused with [U^13^C]-glucose, and patients I and II received no infusion. Line inside of each box represents median, boxes represent interquartile range, whiskers represent minimum and maximum values, and points outside whiskers indicate outliers.

**Figure S7.** Production of ribose 5-phosphate from glucose in cortex and brain tumors. (A) Fractional enrichment of ribose 5-phosphate in tissues from orthotopic GBM bearing mice (n=7) infused with [U^13^C]-glucose. (B) Normalized (to plasma m+6 glucose on a per-patient basis) enrichment of ribose 5-phosphate in cortex and tumor tissues from glioma patients infused with [U^13^C]-glucose. Data are shown as average ± standard deviation. ns, not significant.

**Figure S8.** Glucose-derived production of inosylate and adenylate purine metabolites in human cortex and glioma. Normalized (to plasma m+6 glucose on a per-patient basis) enrichment of (A) IMP, (B) inosine, (C) AMP, and (D) ADP. Data are shown as average ± standard deviation. *p<0.05. **p<0.01. ***p<0.001. ****p<0.0001. ns, not significant. Abbreviations: IMP, inosine monophosphate; AMP, adenosine monophosphate; ADP, adenosine diphosphate.

**Figure S9.** Glucose-derived production of guanylate purine metabolites in human cortex and glioma. Normalized (to plasma m+6 glucose on a per-patient basis) enrichment of (A) guanosine, (B) GMP, and (C) GDP. Data are shown as average ± standard deviation. *p<0.05. **p<0.01. ***p<0.001. ****p<0.0001. Abbreviations: GMP, guanosine monophosphate; GDP, guanosine diphosphate.

**Figure S10.** Glucose-derived production of pyrimidine metabolites in human cortex and glioma. Normalized (to plasma m+6 glucose on a per-patient basis) enrichment of (A) UMP and (B) dTDP. Data are shown as average ± standard deviation. *p<0.05. **p<0.01. ***p<0.001. ****p<0.0001. Abbreviations: UMP, uridine monophosphate; dTDP, deoxythymidine diphosphate.

**Figure S11.** Glucose-derived production of NAD in cortex and GBM. (A) Schematic of carbon incorporation into NAD and NADH. (B) ^13^C enrichment of NAD and NADH in cortex and GBM tissue from orthotopic GBM-bearing mice infused with [U^13^C]-glucose. Data are shown as average ± standard deviation. N=7 mice. **p<0.01. Abbreviations: NAM, nicotinamide; ATP, adenosine triphosphate; NAD, oxidized nicotinamide adenine dinucleotide; NADH, reduced nicotinamide adenine dinucleotide.

**Figure S12.** Time-course incorporation of [U^13^C]-glucose carbons into nucleotide metabolites in cortex and GBM. (A) Time-dependent enrichment profiles of purine metabolites IMP, GDP, guanosine, AMP, and inosine in cortex and GBM in orthotopic brain tumor bearing mice infused with [U^13^C]-glucose. (B) Time-dependent enrichment of pyrimidine metabolites UMP and uridine in cortex and GBM in orthotopic brain tumor bearing mice infused with [U^13^C]-glucose. (C) Relative abundance of nucleotide species in cortical and GBM tissues from orthotopic tumor bearing mice. Data are shown as average ± standard deviation, n=3-9 samples from 1-3 mice per time point.

**Figure S13.** Pathway based framework for *in vivo* modeling of nucleotide synthesis. Biochemical interconversions and model boundaries used for metabolic modeling of (A) purine synthesis and (B) pyrimidine synthesis.

**Figure S14.** Decreased glucose-derived TCA cycle activity in GBM compared to cortex. Isotopologue abundance levels of citrate in cortex (A) and GBM (B) tissues at indicated time points from orthotopic GBM bearing mice infused with [U^13^C]-glucose. (C) Percent change in indicated citrate isotopologues from 120 to 240 min. Error bars are propagated from uncertainty in t=120 and t=240 min datapoints in (A) and (B). Isotopologue abundance levels of succinate in cortex (D) and GBM (E) in mice infused with [U^13^C]-glucose. (F) Percent change in indicated succinate isotopologues from 120 to 240 min. Error bars are propagated from uncertainty in t=120 and t=240 min datapoints in (D) and (E). Data are shown as average ± standard deviation, n=6-9 samples from 2-3 mice per time point.

**Figure S15.** Time-dependent changes in purine synthesis activity after RT in cortex and GBM. (A) Isotope enrichment and (B) relative abundance of purine metabolites IMP, GDP, guanosine, AMP, and inosine in cortex and GBM in orthotopic brain tumor bearing mice treated with a single dose of cranial RT (8 Gy) and then immediately infused with [U^13^C]-glucose. Data are shown as average ± standard deviation, n=3-9 samples from 1-3 mice per time point.

**Figure S16.** Metabolic reaction fluxes after RT in GBM and cortex. Flux values were approximated using time-course enrichment profiles from mice infused with [U^13^C]-glucose as described in supplementary methods.

**Figure S17.** Physiological glucose-derived serine production patterns reported in previous studies. (A) Isotopic enrichment of serine in brain tumors from patients infused with [U^13^C]-glucose were plotted from supplemental data reported by Courtney *et al*. (B-D) Isotopic enrichment of serine in different pediatric tumor types including (B) pediatric neuroblastoma, (C) pediatric sarcoma, and (D) a variety of other miscellaneous pediatric cancers was previously reported as supplemental information by Johnston *et al*. and plotted here. (E) The work of Courtney *et al*. includes supplemental information reporting enrichment of serine in clear cell renal cell carcinoma (CCRC) and adjacent kidney tissue. (F) The work of Hui *et al*. includes ^13^C serine labeling information in multiple tissue types from mice infused with [U^13^C]-glucose. Data are shown as average ± standard deviation.

**Figure S18.** Different serine sources in human cortex and brain cancers. (A) Statistical analysis of M+1 and M+3 serine enrichment confirms higher M+1 serine and lower M+3 serine in brain cancer compared to the cortex. Data are shown as average ± standard deviation. *p<0.05. **p<0.01. ***p<0.001. ****p<0.0001. (B) Ratio of contribution of *de novo* serine synthesis to serine salvage in human cortex and brain cancers. A value higher than 1 indicates that *de novo* synthesis is the predominant source of labeled serine, while a value lower than 1 indicates that serine salvage is the predominant source of labeled serine. The error bars represent 95% confidence intervals.

**Figure S19.** Plasma serine levels and tissue metabolites in orthotopic GBM bearing mice on a serine/glycine-restricted diet. (A) Levels of serine in plasma from orthotopic GBM bearing mice fed either a control diet or serine/glycine-restricted diet, n=5 mice per group. (B) Principal component analysis of cortex and GBM tissues from mice on a control diet (n=4) or serine/glycine-restricted diet (n=5) in panel A. (C) Individual metabolite levels in cortical tissue from mice in panel A. One mouse in the control diet group in panel A was found to have no detectable tumor and was therefore excluded from analyses in panels B and C.
