## Supplemental Figures for "Rewiring of cortical glucose metabolism fuels human brain cancer growth"

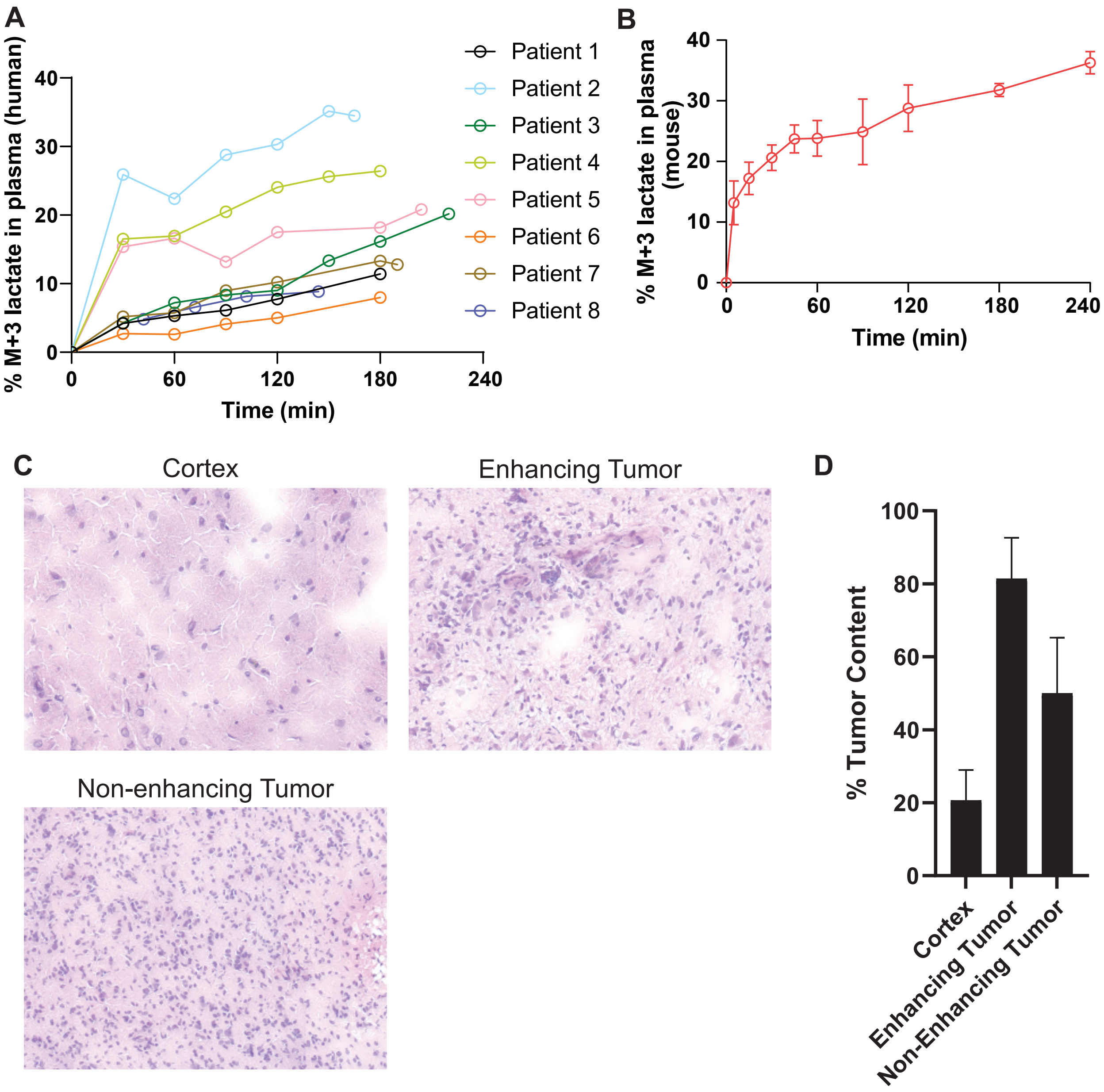

A
% M+3 lactate in plasma (human)
Time (min)
Patient 1
Patient 2
Patient 3
Patient 4
Patient 5
Patient 6
Patient 7
Patient 8
B
% M+3 lactate in plasma (mouse)
Time (min)
C
Cortex
Enhancing Tumor
Non-enhancing Tumor
D
% Tumor Content
Cortex
Enhancing Tumor
Non-Enhancing Tumor


### Fig S1

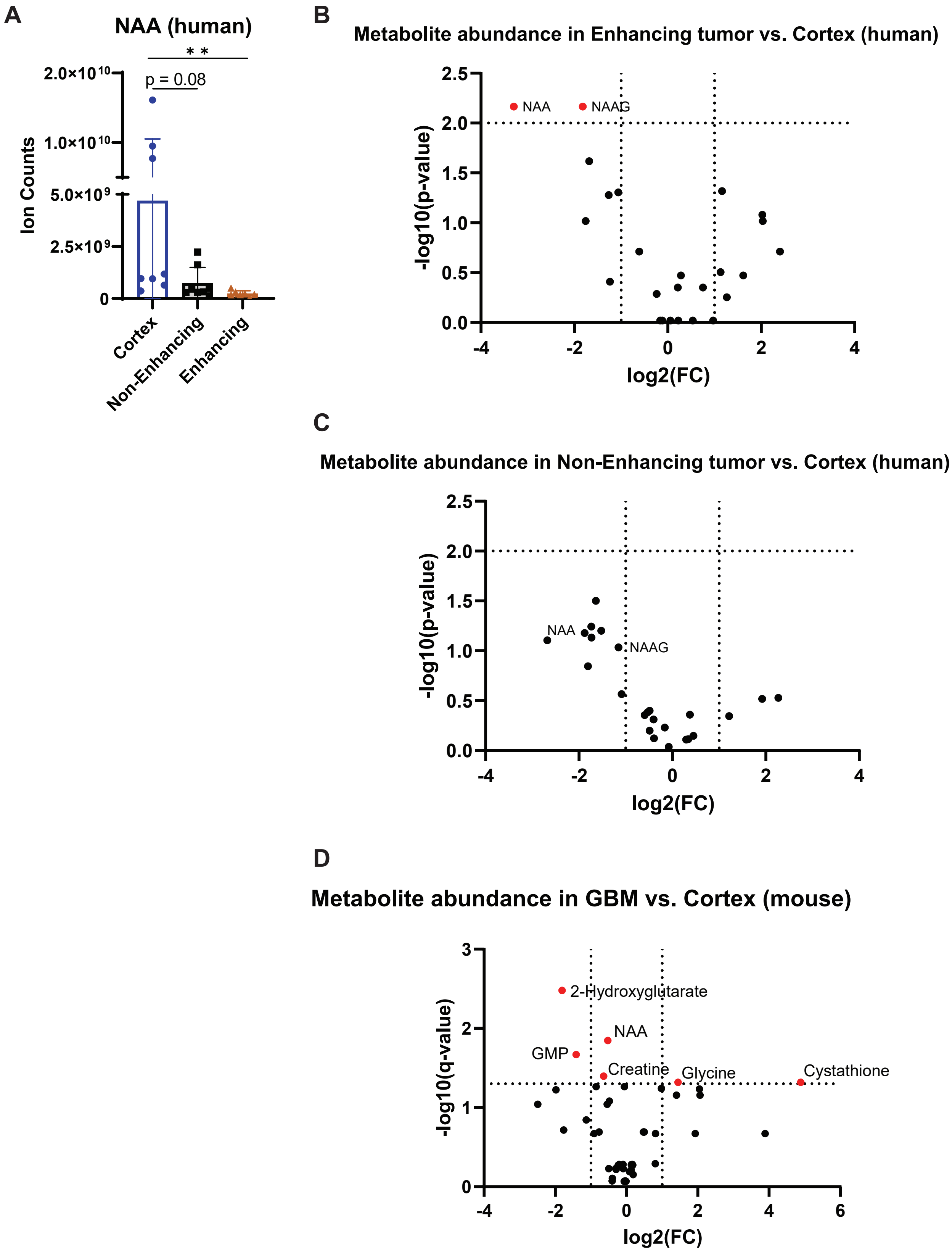
A
NAA (human)
* *
p = 0.08
2.0×10^10
1.0×10^10
5.0×10^9
2.5×10^9
0
Ion Counts
Cortex
Non-Enhancing
Enhancing
B
Metabolite abundance in Enhancing tumor vs. Cortex (human)
NAA
NAAG
-log10(p-value)
log2(FC)
C
Metabolite abundance in Non-Enhancing tumor vs. Cortex (human)
NAA
NAAG
-log10(p-value)
log2(FC)
D
Metabolite abundance in GBM vs. Cortex (mouse)
2-Hydroxyglutarate
NAA
GMP
Creatine
Glycine
Cystathione
-log10(q-value)
log2(FC)

Fig S2

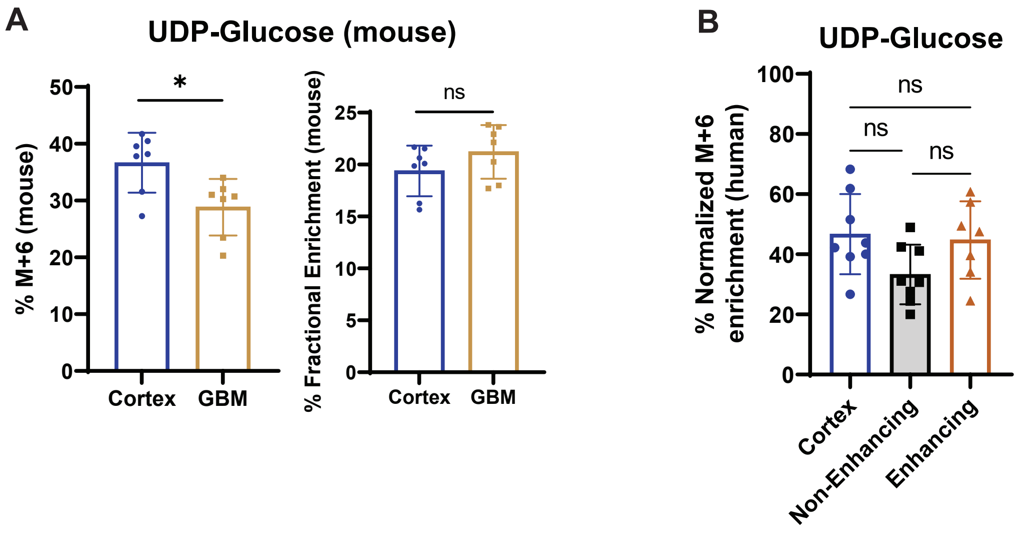
A
UDP-Glucose (mouse)
% M+6 (mouse)
50
40
30
20
10
0
*
Cortex
GBM
% Fractional Enrichment (mouse)
25
20
15
10
5
0
ns
Cortex
GBM
B
UDP-Glucose
% Normalized M+6 enrichment (human)
100
80
60
40
20
0
ns
ns
ns
Cortex
Non-Enhancing
Enhancing

Fig S3

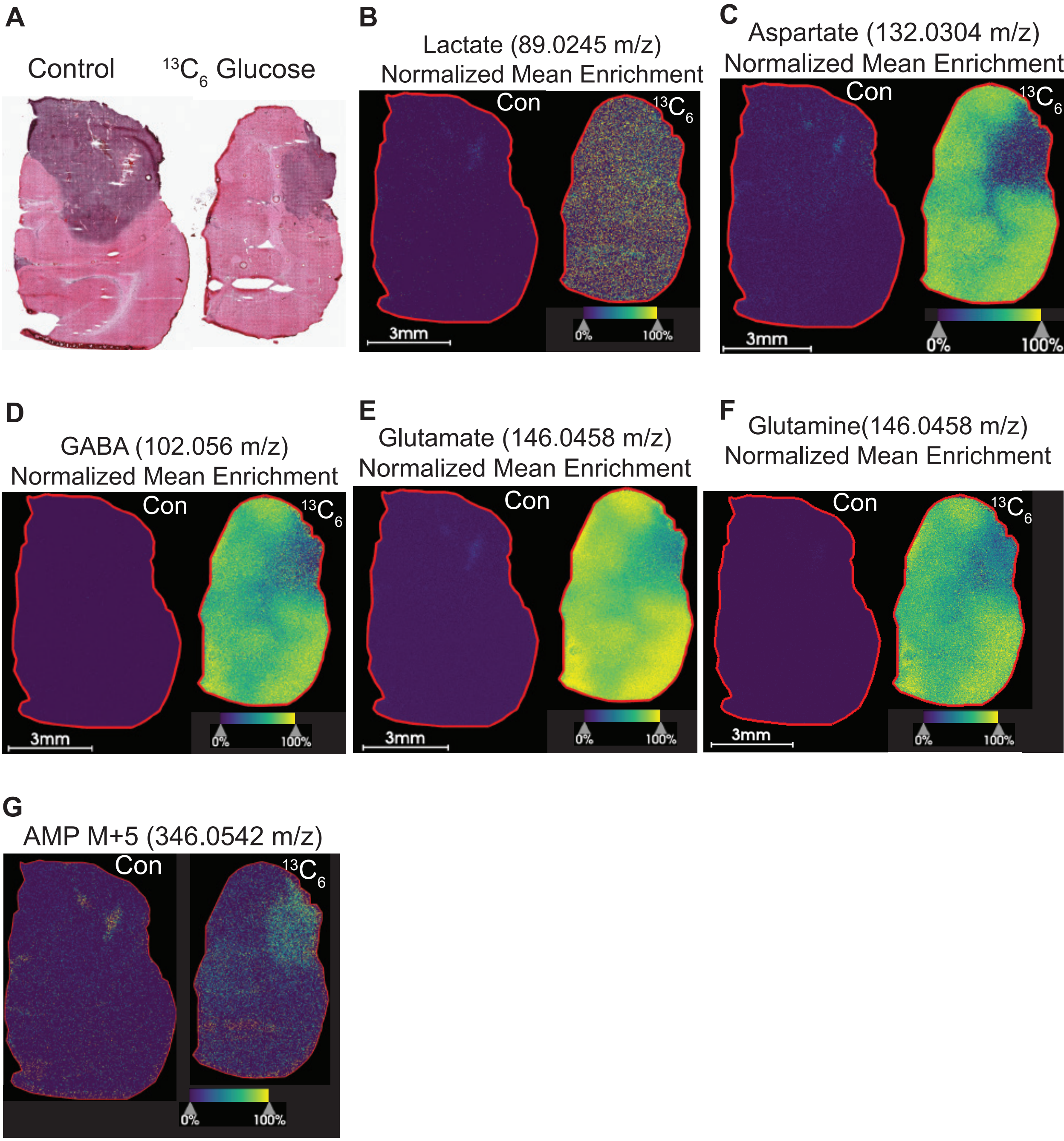
A
Control
13C6 Glucose
B
Lactate (89.0245 m/z)
Normalized Mean Enrichment
Con
13C6
3mm
0%
100%
C
Aspartate (132.0304 m/z)
Normalized Mean Enrichment
Con
13C6
3mm
0%
100%
D
GABA (102.056 m/z)
Normalized Mean Enrichment
Con
13C6
3mm
0%
100%
E
Glutamate (146.0458 m/z)
Normalized Mean Enrichment
Con
3mm
0%
100%
F
Glutamine(146.0458 m/z)
Normalized Mean Enrichment
Con
13C6
3mm
0%
100%
G
AMP M+5 (346.0542 m/z)
Con
13C6
0%
100%

Fig S4

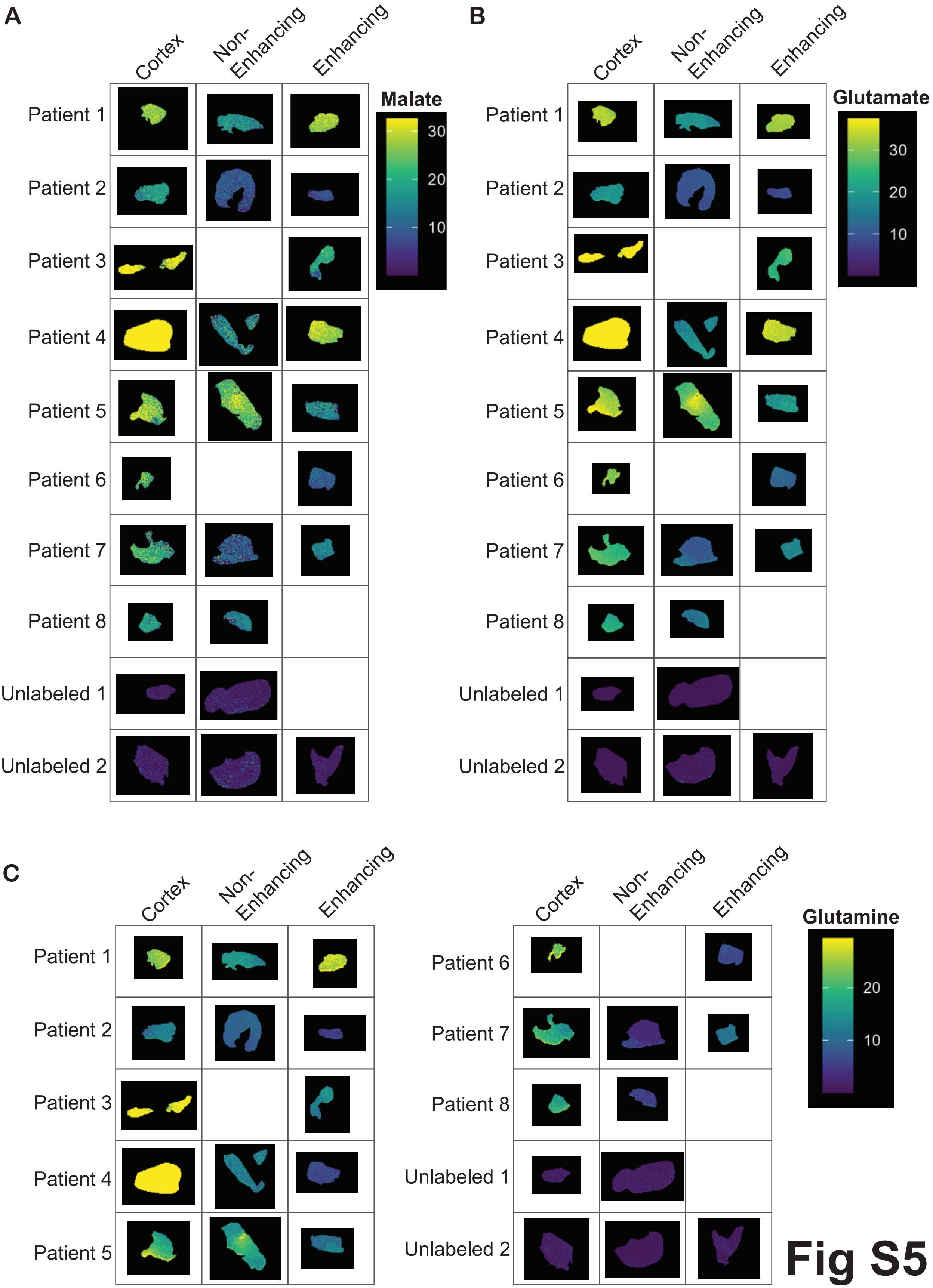
A
Cortex
Non-Enhancing
Enhancing
Patient 1
Patient 2
Patient 3
Patient 4
Patient 5
Patient 6
Patient 7
Patient 8
Unlabeled 1
Unlabeled 2
Malate
30
20
10
B
Cortex
Non-Enhancing
Enhancing
Patient 1
Patient 2
Patient 3
Patient 4
Patient 5
Patient 6
Patient 7
Patient 8
Unlabeled 1
Unlabeled 2
Glutamate
30
20
10
C
Cortex
Non-Enhancing
Enhancing
Patient 1
Patient 2
Patient 3
Patient 4
Patient 5
Cortex
Non-Enhancing
Enhancing
Patient 6
Patient 7
Patient 8
Unlabeled 1
Unlabeled 2
Glutamine
20
10

Fig S5

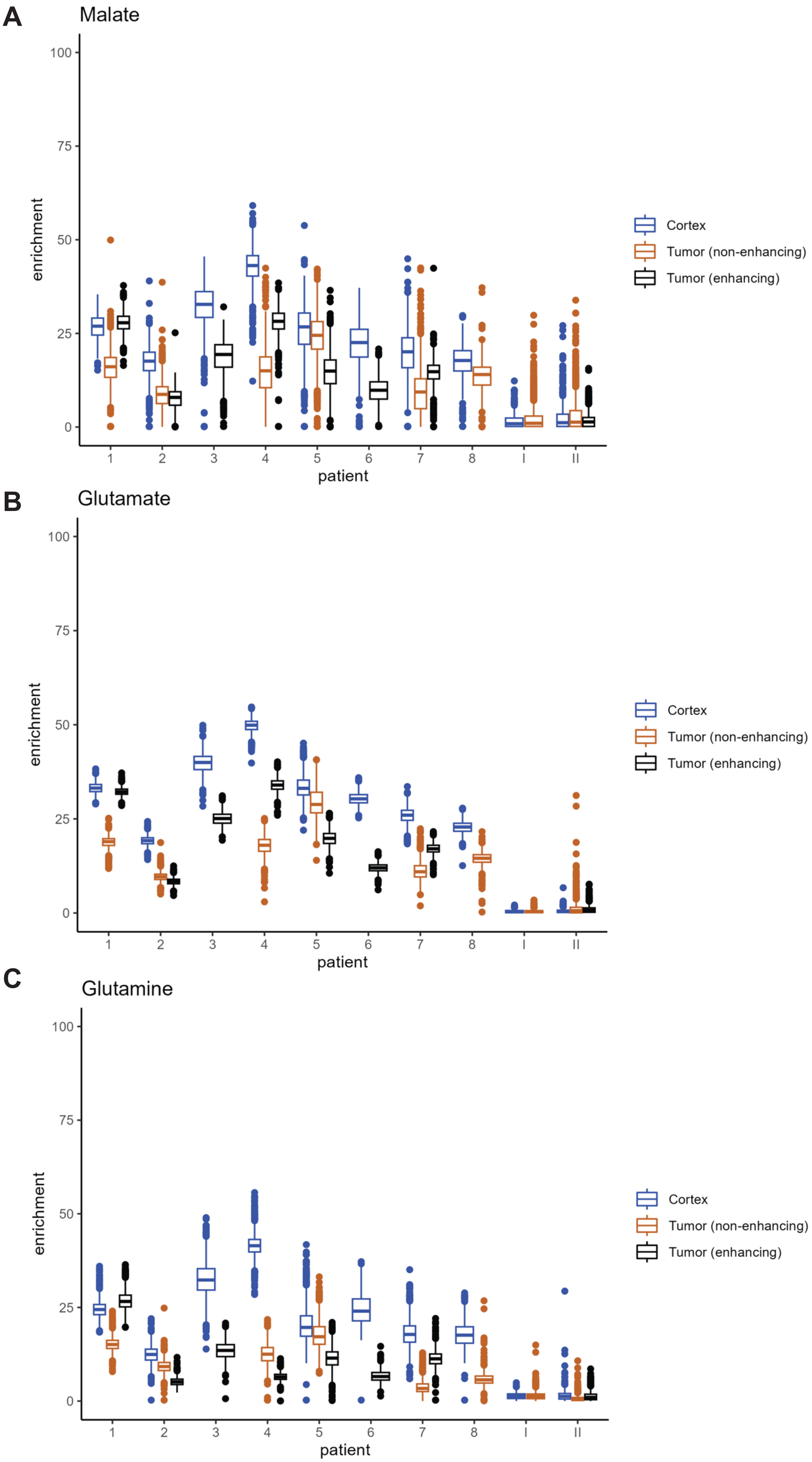

A
Malate
enrichment
patient
B
Glutamate
C
Glutamine
Cortex
Tumor (non-enhancing)
Tumor (enhancing)


Fig S6

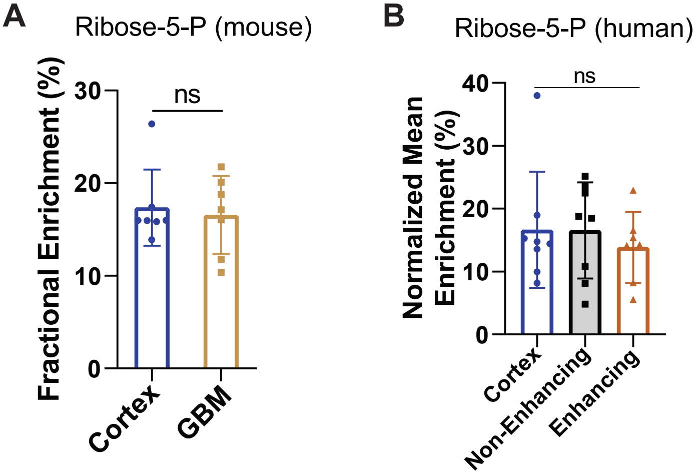
A
Ribose-5-P (mouse)
Fractional Enrichment (%)
30
20
10
0
ns
Cortex
GBM
B
Ribose-5-P (human)
Normalized Mean Enrichment (%)
40
30
20
10
0
ns
Cortex
Non-Enhancing
Enhancing

Fig S7

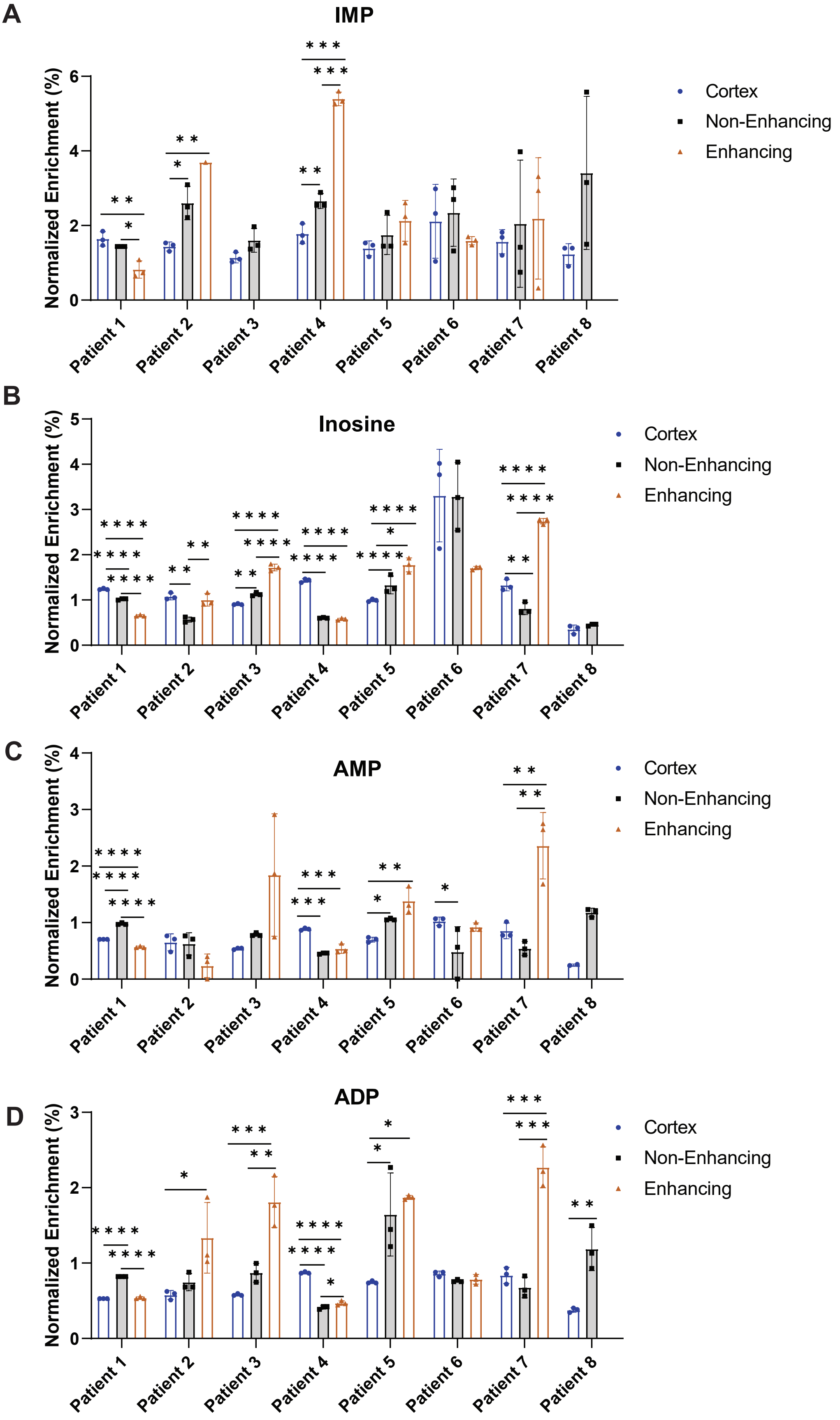
A
IMP
Normalized Enrichment (%)
0
2
4
6
Cortex
Non-Enhancing
Enhancing
Patient 1
Patient 2
Patient 3
Patient 4
Patient 5
Patient 6
Patient 7
Patient 8
B
Inosine
Normalized Enrichment (%)
0
1
2
3
4
5
Cortex
Non-Enhancing
Enhancing
Patient 1
Patient 2
Patient 3
Patient 4
Patient 5
Patient 6
Patient 7
Patient 8
C
AMP
Normalized Enrichment (%)
0
1
2
3
4
Cortex
Non-Enhancing
Enhancing
Patient 1
Patient 2
Patient 3
Patient 4
Patient 5
Patient 6
Patient 7
Patient 8
D
ADP
Normalized Enrichment (%)
0
1
2
3
Cortex
Non-Enhancing
Enhancing
Patient 1
Patient 2
Patient 3
Patient 4
Patient 5
Patient 6
Patient 7
Patient 8

Fig S8

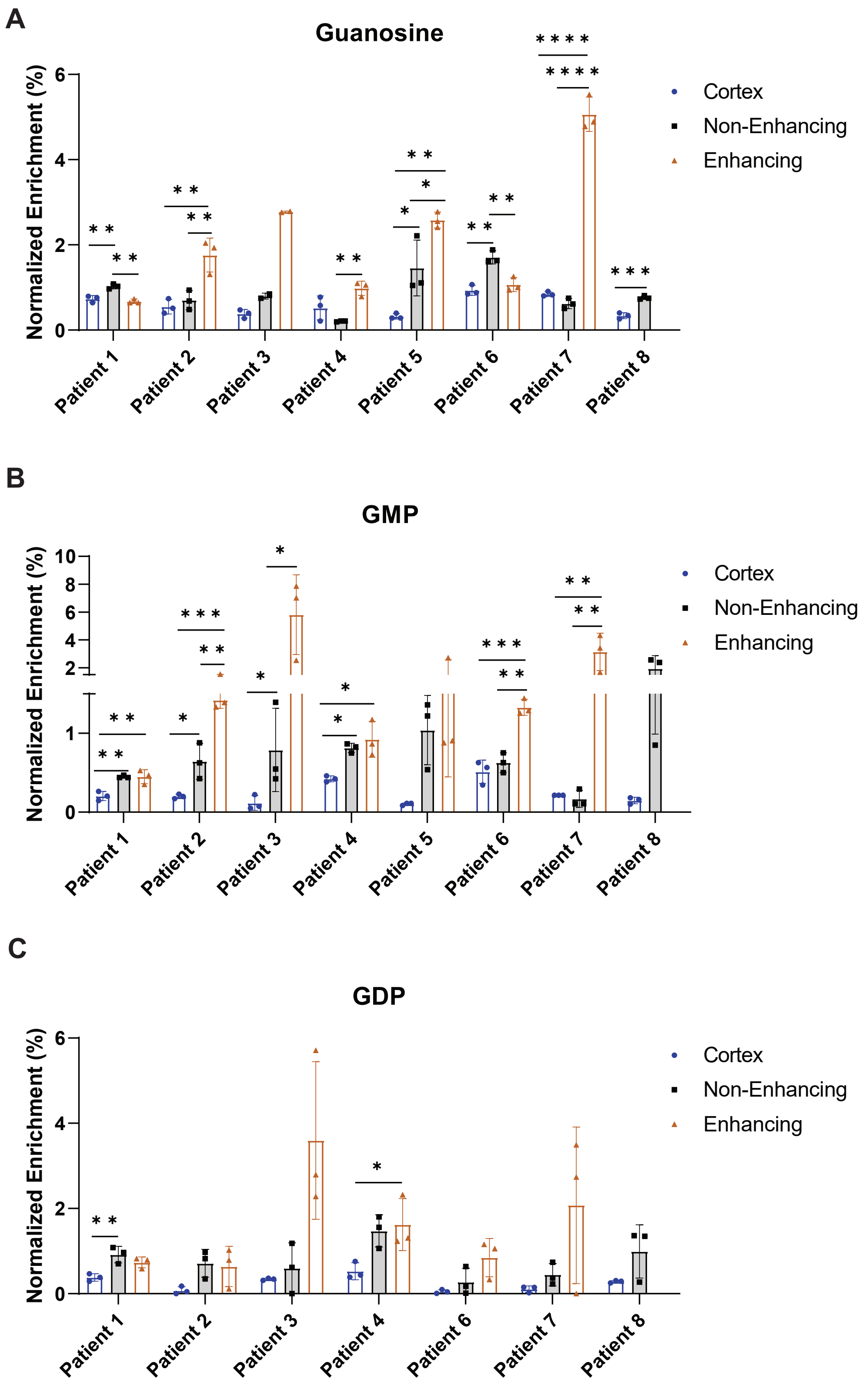
A
Guanosine
Normalized Enrichment (%)
Cortex
Non-Enhancing
Enhancing
Patient 1
Patient 2
Patient 3
Patient 4
Patient 5
Patient 6
Patient 7
Patient 8
B
GMP
Normalized Enrichment (%)
Cortex
Non-Enhancing
Enhancing
Patient 1
Patient 2
Patient 3
Patient 4
Patient 5
Patient 6
Patient 7
Patient 8
C
GDP
Normalized Enrichment (%)
Cortex
Non-Enhancing
Enhancing
Patient 1
Patient 2
Patient 3
Patient 4
Patient 6
Patient 7
Patient 8

Fig S9

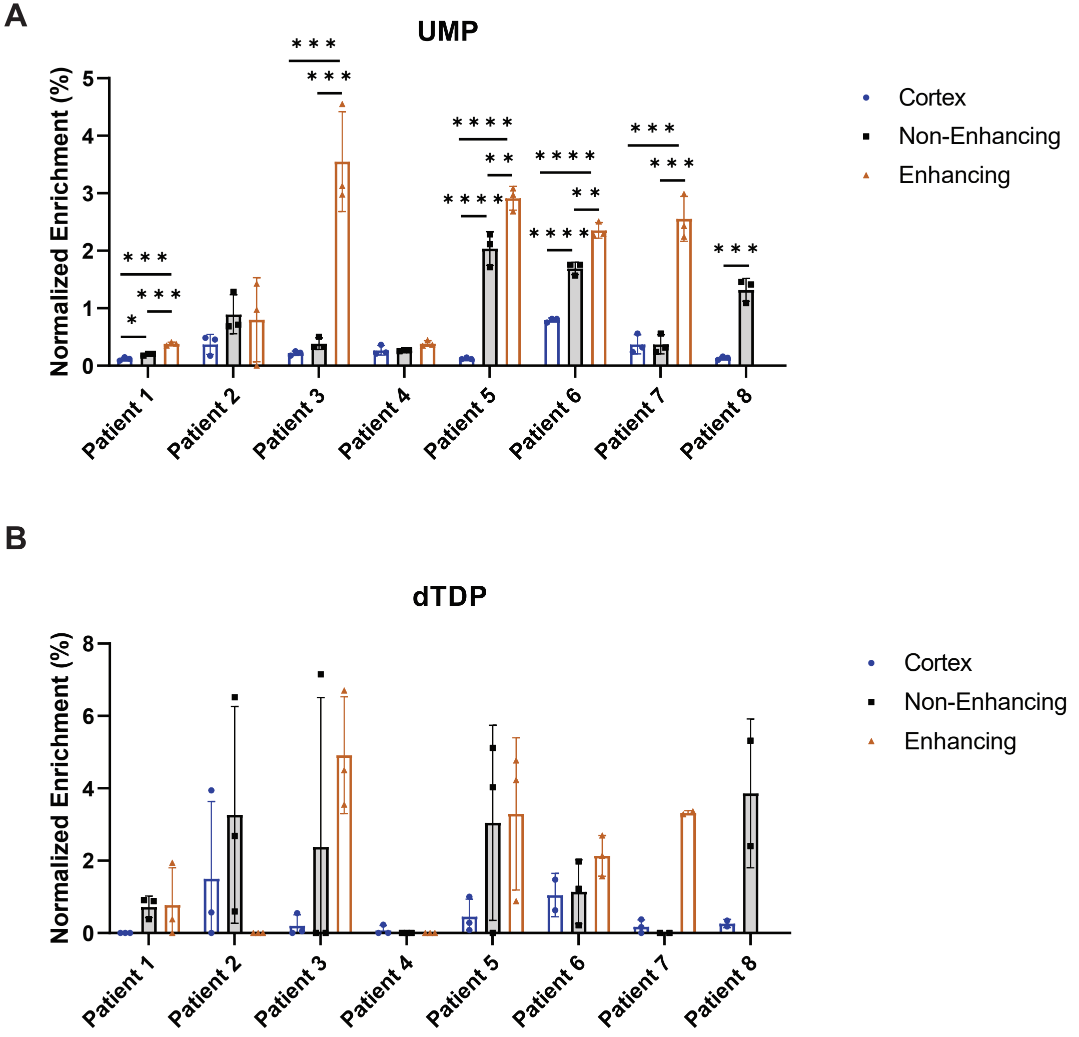

A
UMP
Normalized Enrichment (%)
Cortex
Non-Enhancing
Enhancing
Patient 1
Patient 2
Patient 3
Patient 4
Patient 5
Patient 6
Patient 7
Patient 8
B
dTDP
Normalized Enrichment (%)
Cortex
Non-Enhancing
Enhancing
Patient 1
Patient 2
Patient 3
Patient 4
Patient 5
Patient 6
Patient 7
Patient 8


**Fig S10**

**A**

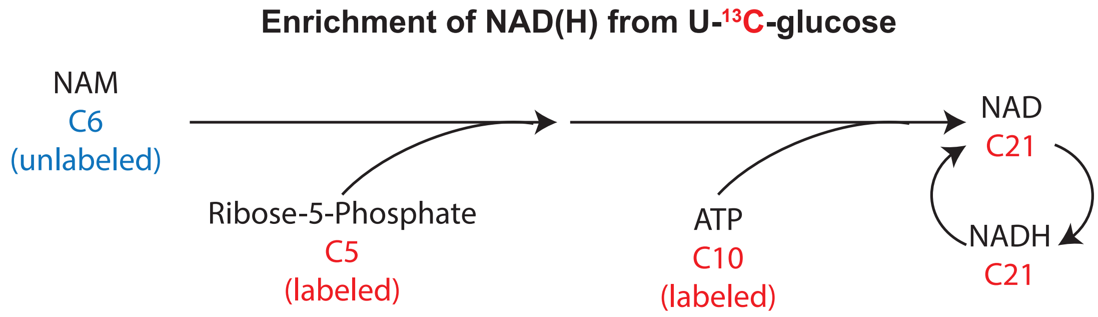
Enrichment of NAD(H) from U-13C-glucose
NAM
C6
(unlabeled)
Ribose-5-Phosphate
C5
(labeled)
ATP
C10
(labeled)
NAD
C21
NADH
C21

**B**

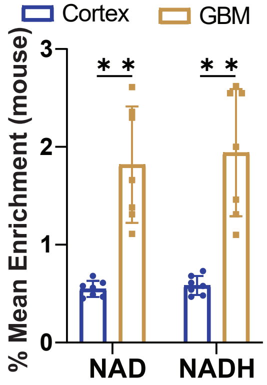
Cortex
GBM
% Mean Enrichment (mouse)
3
2
1
0
* *
* *
NAD
NADH

### Fig S11

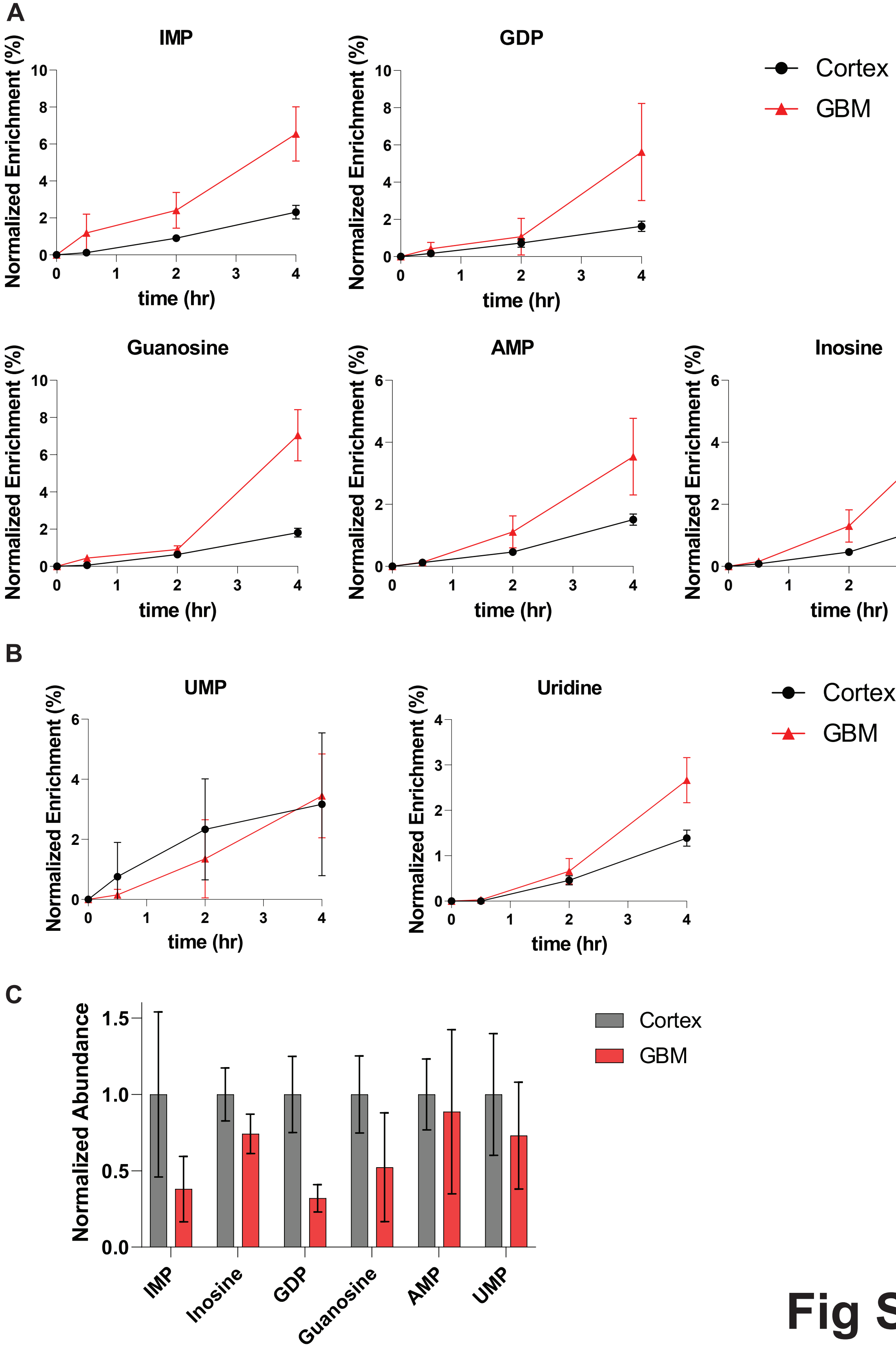
A
IMP
GDP
Cortex
GBM
Guanosine
AMP
Inosine
Normalized Enrichment (%)
time (hr)
B
UMP
Uridine
Cortex
GBM
C
Normalized Abundance
Cortex
GBM
IMP
Inosine
GDP
Guanosine
AMP
UMP

Fig S12

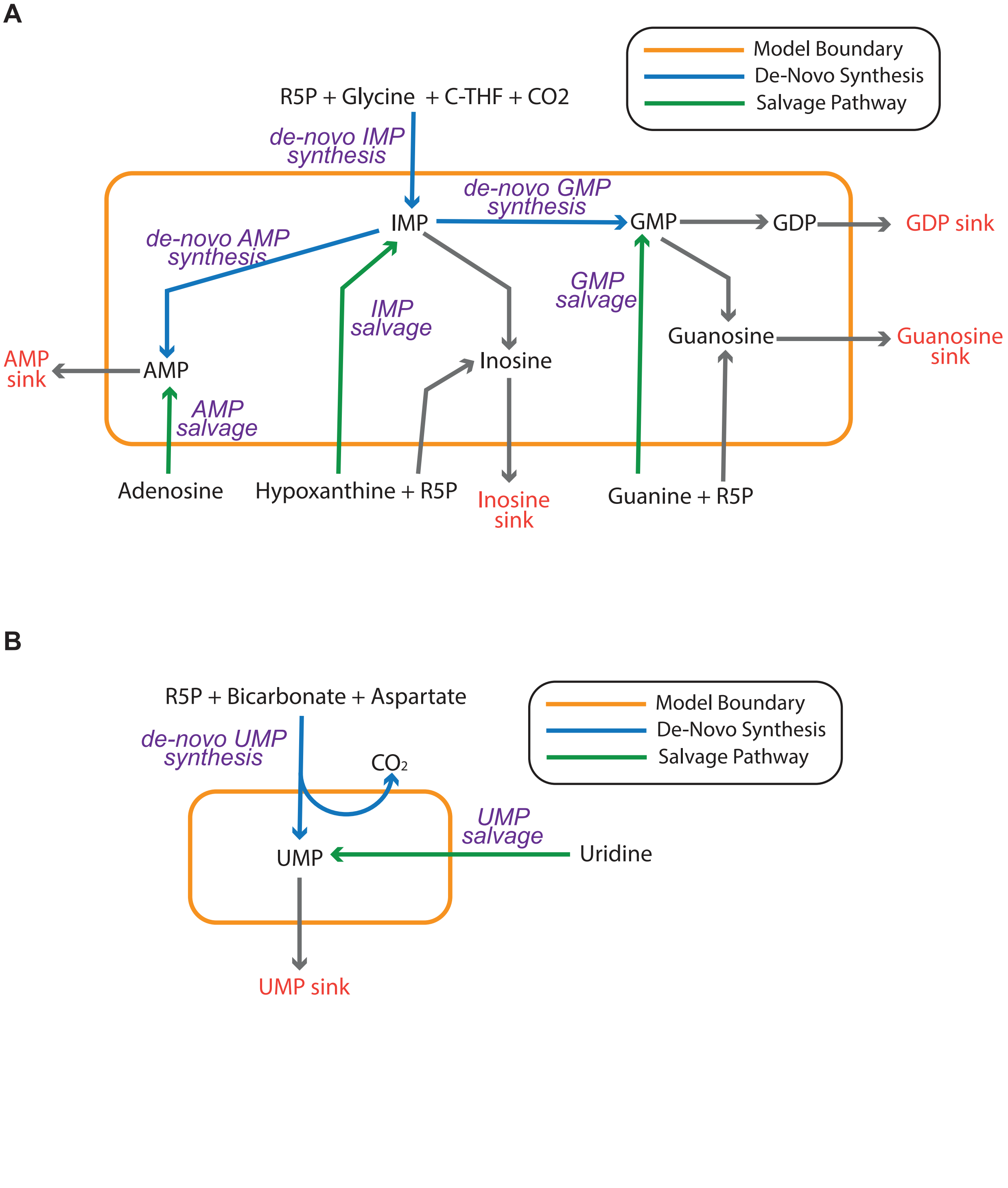
A
Model Boundary
De-Novo Synthesis
Salvage Pathway
R5P + Glycine + C-THF + CO2
de-novo IMP synthesis
de-novo GMP synthesis
IMP
GMP
GDP
GDP sink
de-novo AMP synthesis
GMP salvage
IMP salvage
Guanosine
Guanosine sink
AMP sink
AMP
Inosine
AMP salvage
Adenosine
Hypoxanthine + R5P
Inosine sink
Guanine + R5P
B
R5P + Bicarbonate + Aspartate
Model Boundary
De-Novo Synthesis
Salvage Pathway
de-novo UMP synthesis
CO2
UMP salvage
UMP
Uridine
UMP sink

Fig S13

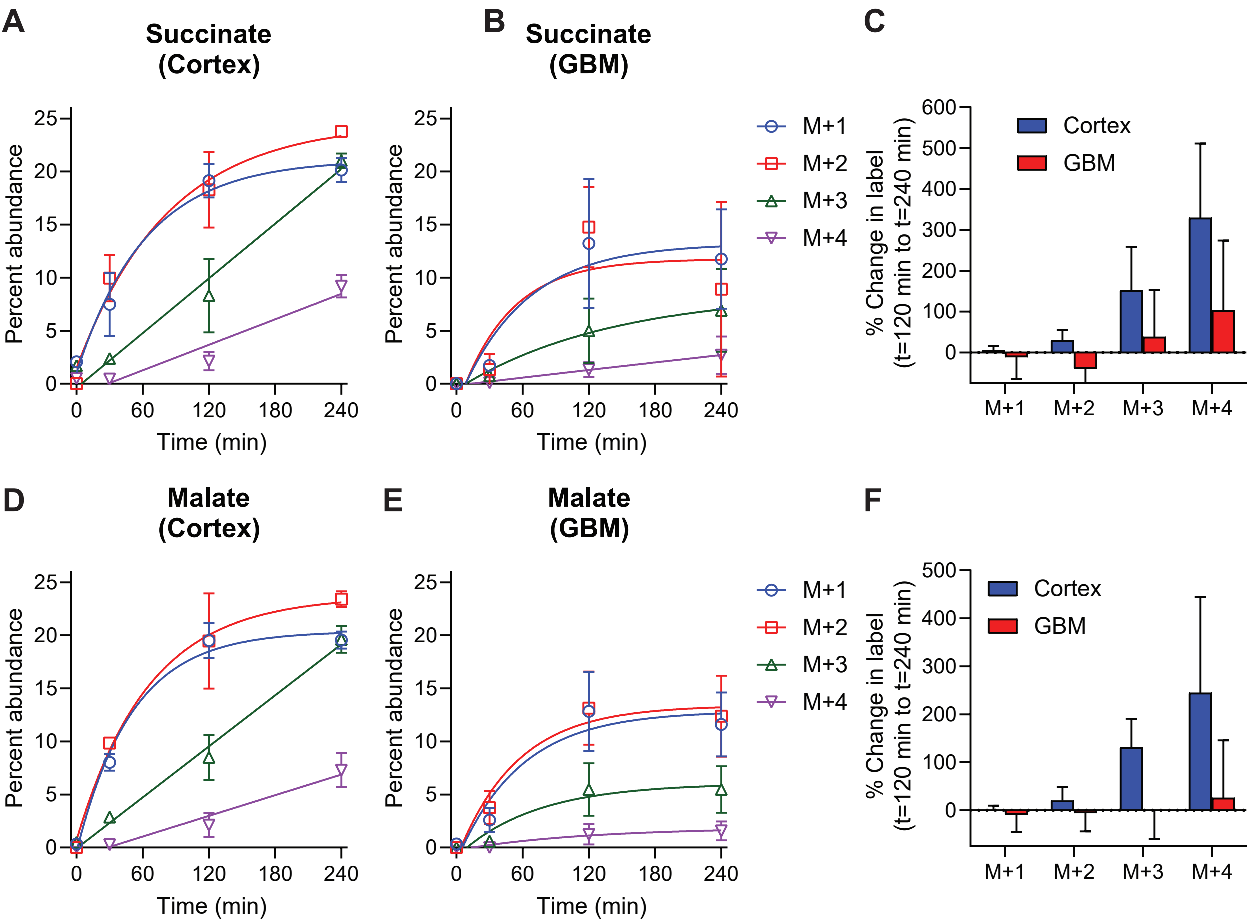
A
Succinate (Cortex)
B
Succinate (GBM)
C
D
Malate (Cortex)
E
Malate (GBM)
F
Percent abundance
Time (min)
M+1
M+2
M+3
M+4
Cortex
GBM
% Change in label (t=120 min to t=240 min)

### Fig S14

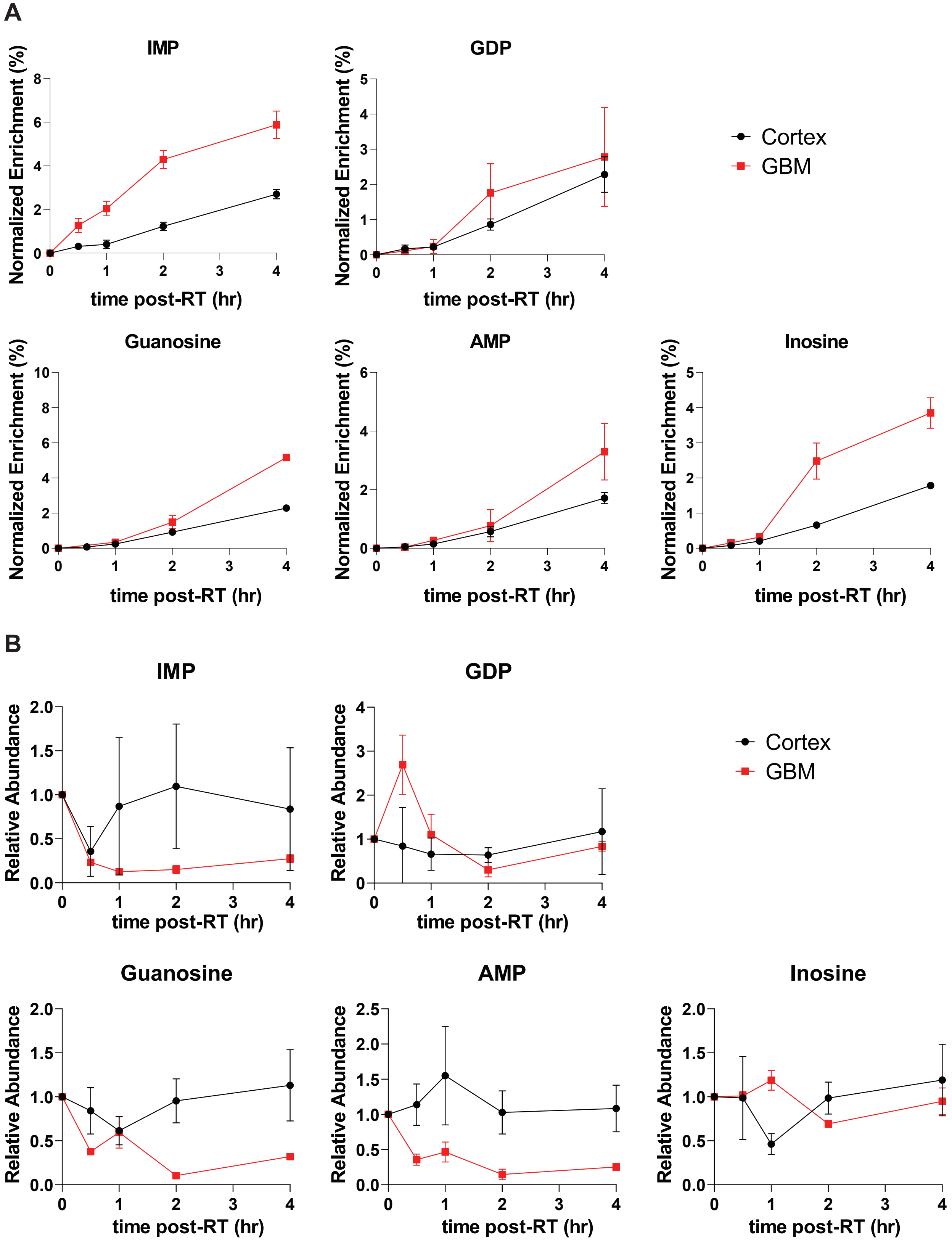
A
IMP
GDP
Normalized Enrichment (%)
time post-RT (hr)
Cortex
GBM
Guanosine
AMP
Inosine
B
Relative Abundance

Fig S15

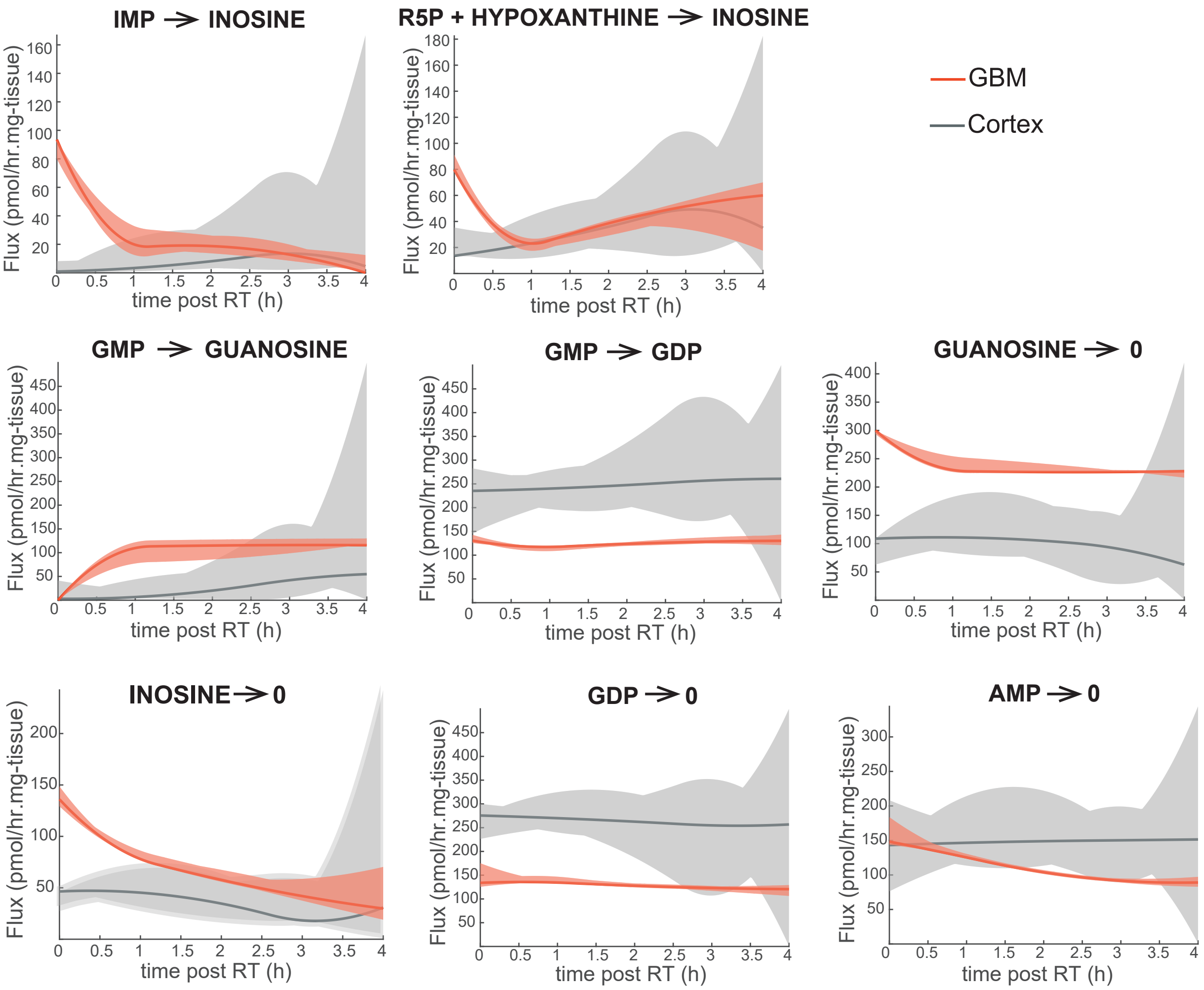
IMP → INOSINE
R5P + HYPOXANTHINE → INOSINE
GBM
Cortex
GMP → GUANOSINE
GMP → GDP
GUANOSINE → 0
INOSINE → 0
GDP → 0
AMP → 0
Flux (pmol/hr.mg-tissue)
time post RT (h)

Fig S16

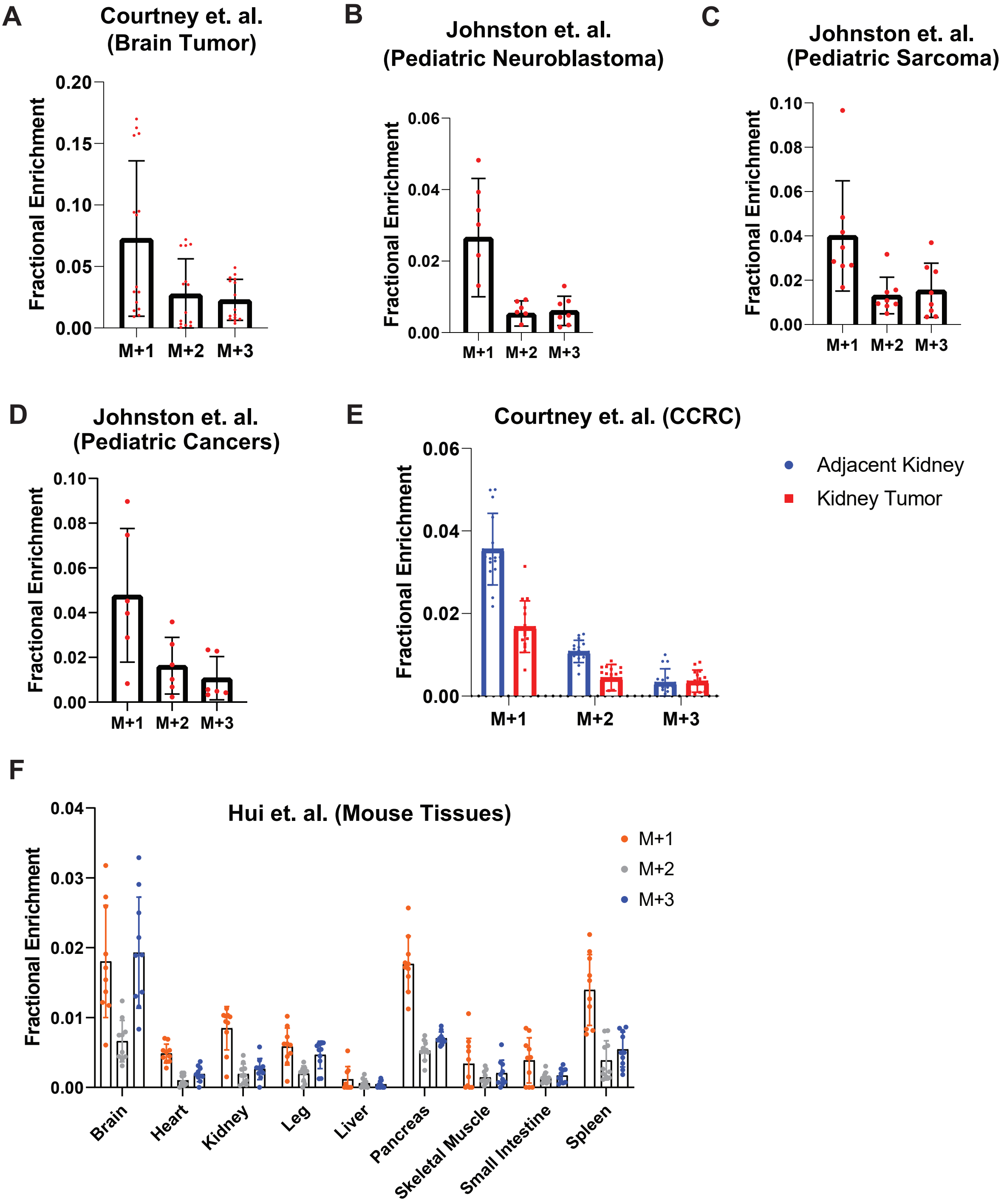

A
Courtney et. al.
(Brain Tumor)
Fractional Enrichment
0.20
0.15
0.10
0.05
0.00
M+1
M+2
M+3
B
Johnston et. al.
(Pediatric Neuroblastoma)
Fractional Enrichment
0.06
0.04
0.02
0.00
M+1
M+2
M+3
C
Johnston et. al.
(Pediatric Sarcoma)
Fractional Enrichment
0.10
0.08
0.06
0.04
0.02
0.00
M+1
M+2
M+3
D
Johnston et. al.
(Pediatric Cancers)
Fractional Enrichment
0.10
0.08
0.06
0.04
0.02
0.00
M+1
M+2
M+3
E
Courtney et. al. (CCRC)
Fractional Enrichment
0.06
0.04
0.02
0.00
M+1
M+2
M+3
Adjacent Kidney
Kidney Tumor
F
Hui et. al. (Mouse Tissues)
Fractional Enrichment
0.04
0.03
0.02
0.01
0.00
M+1
M+2
M+3
Brain
Heart
Kidney
Leg
Liver
Pancreas
Skeletal Muscle
Small Intestine
Spleen


Fig S17

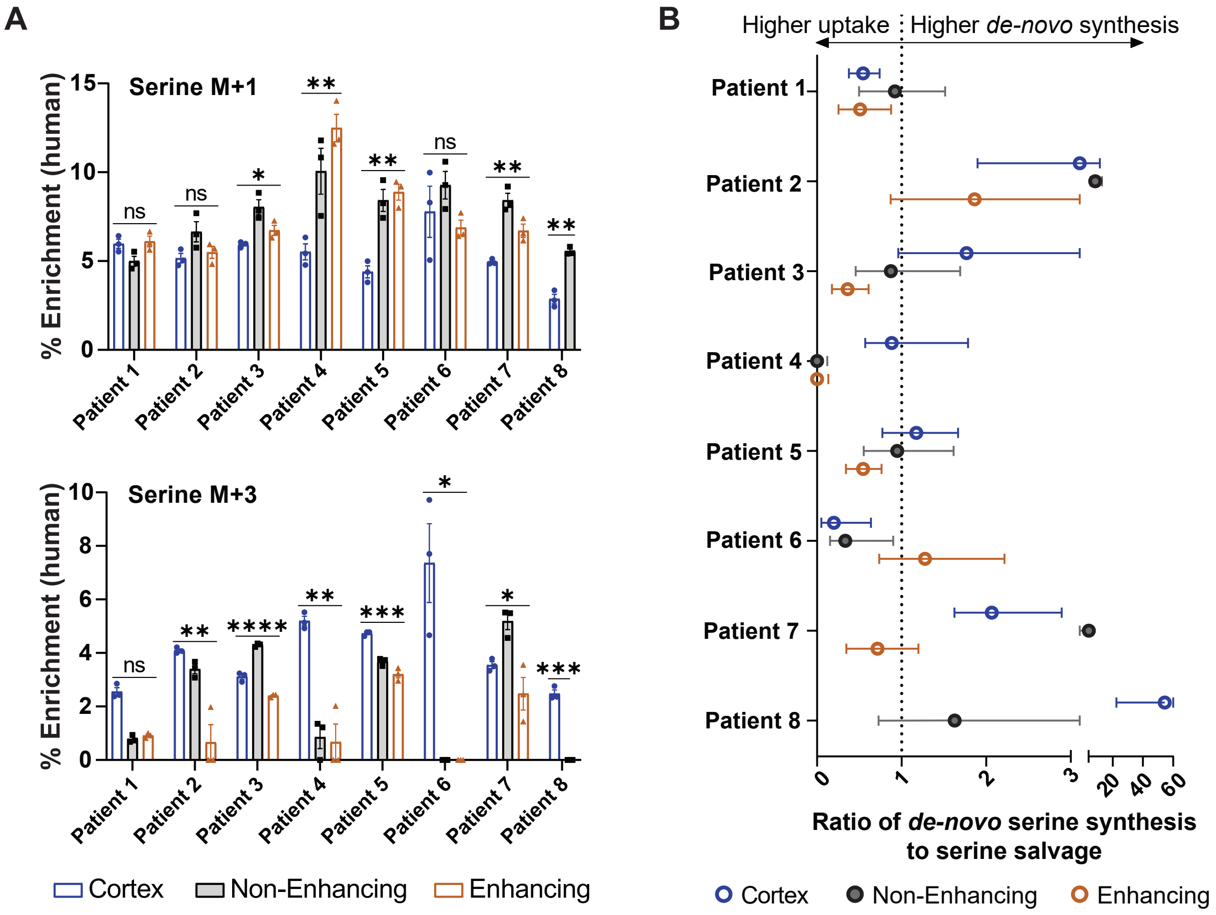
A
Serine M+1
% Enrichment (human)
Serine M+3
Patient 1
Patient 2
Patient 3
Patient 4
Patient 5
Patient 6
Patient 7
Patient 8
Cortex
Non-Enhancing
Enhancing
B
Higher uptake
Higher de-novo synthesis
Ratio of de-novo serine synthesis to serine salvage


### Fig S18

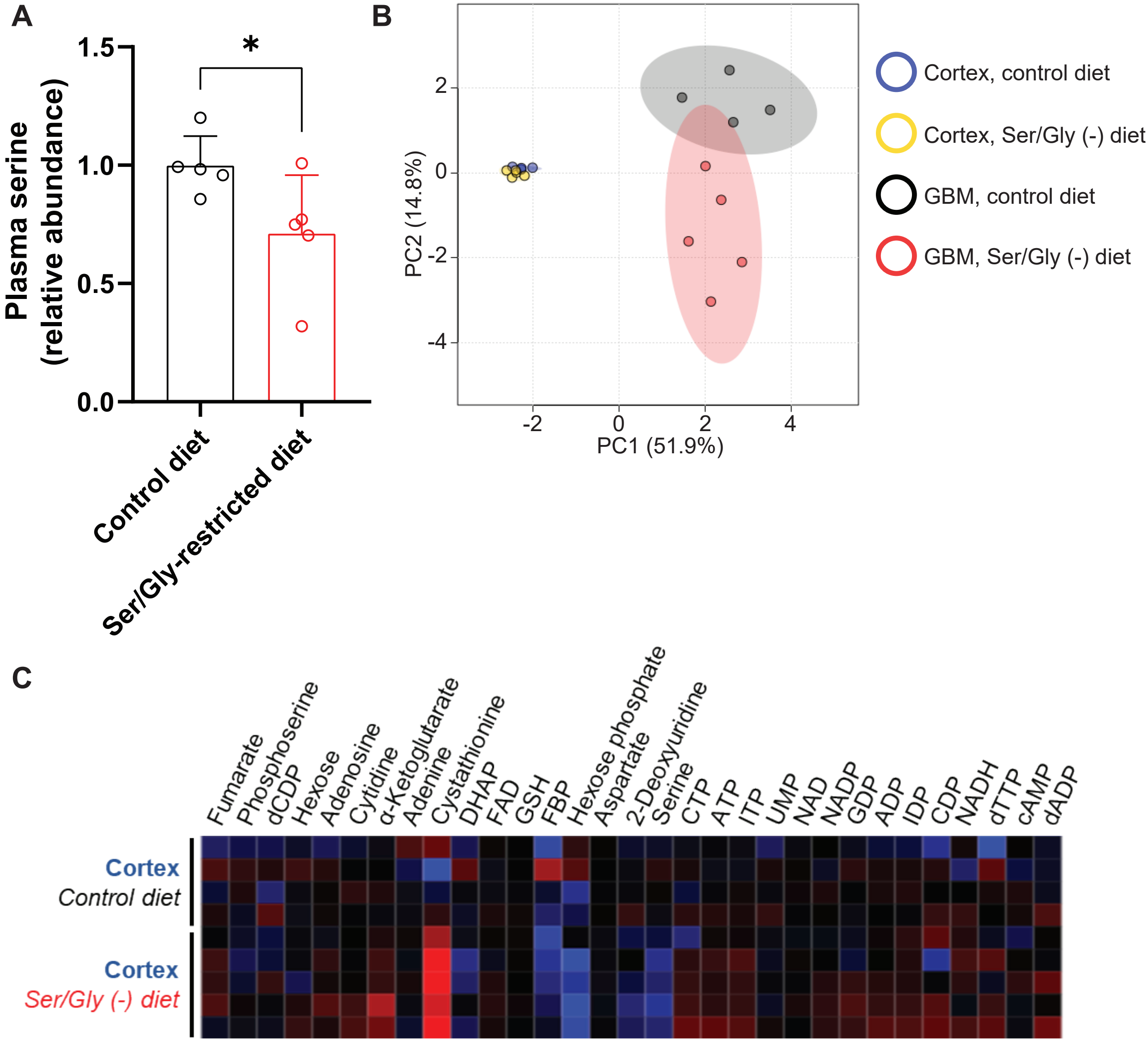
A
Plasma serine (relative abundance)
1.5
1.0
0.5
0.0
*
Control diet
Ser/Gly-restricted diet
B
PC2 (14.8%)
PC1 (51.9%)
Cortex, control diet
Cortex, Ser/Gly (-) diet
GBM, control diet
GBM, Ser/Gly (-) diet
C
Fumarate
Phosphoserine
dCDP
Hexose
Adenosine
Cytidine
α-Ketoglutarate
Adenine
Cystathionine
DHAP
FAD
GSH
FBP
Hexose phosphate
Aspartate
2-Deoxyuridine
Serine
CTP
ATP
ITP
UMP
NAD
NADP
GDP
ADP
IDP
CDP
NADH
dTTP
cAMP
dADP
Cortex
Control diet
Cortex
Ser/Gly (-) diet

### Fig S19
